# Examining the Clinical Utility of Deep Learning Models for Perivascular Space Segmentation in Alzheimer’s Disease

**DOI:** 10.64898/2026.09.27.26363725

**Authors:** Serena Tang, Kyan Younes, Eesha Penukonda, Isabella Hausle, Pamela Thropp, Janine M. Lupo, Duygu Tosun, the Alzheimer’s Disease Neuroimaging Initiative

## Abstract

Open-source deep learning models promise scalable quantification of MRI-visible perivascular spaces (PVS), an emerging marker of glymphatic dysfunction in Alzheimer’s disease (AD). Whether these models remain valid when applied to heterogeneous AD cohorts and preserve ground-truth derived biological associations has not been established.

We benchmarked five open-source nnU-Net-based PVS segmentation models (nnU-Net T1, nnU-Net T1+FLAIR, mcPVS-Net, MedNet-PVS, ADNI-PVS-Net) against manual labels in 60 multi-site ADNI-3 participants balanced across cognitively unimpaired, mild cognitive impairment, and AD dementia, and in 12 publicly available scans from the VAscular Lesions Detection and segmentatiOn challenge (VALDO) challenge. We evaluated overall, voxel, object, and lesion-wise Dice; stratified performance by region, diagnosis, scanner, PVS severity, and white matter hyperintensity (WMH) burden; and tested whether model-derived PVS burden reproduced ground-truth derived associations with AD-related biomarkers.

Despite high internal performance, all models showed a specialist-generalist trade-off: the AD-trained ADNI-PVS-Net achieved highest internal performance (overall Dice = 0.58±0.14) but degraded most externally (Dice ≤ 0.11), while MedNet-PVS and mcPVS-Net generalized externally (Dice 0.27-0.30) but systematically misestimated PVS burden. Models were relatively robust across scanner and diagnosis but inconsistent across PVS severity, and some misclassified up to 32% of WMH as PVS. Notably, only AD-trained models reproduced the expected PVS-amyloid biomarker relationship; other models returned a null result, despite comparable or superior Dice scores.

These findings show that internal segmentation accuracy and external generalizability alone do not guarantee downstream inferential validity. Careful model selection and cohort-specific validation against manual labels should precede clinical inference in AD.

## 1. Introduction

Emerging evidence positions perivascular spaces (PVS) as a promising early imaging biomarker in Alzheimer’s disease (AD) (1–8). As a critical component of the brain’s waste clearance system, PVS have been shown to facilitate the clearance of neurocellular waste products including β-amyloid (Aβ) and tau (1,9,10) by providing a conduit for interstitial and cerebral spinal fluid flow along arterioles and capillaries of the surrounding brain tissue (5,11). On clinical magnetic resonance (MR) imaging at field strengths of 3T and lower, visible PVS are often considered “enlarged” PVS, suggesting localized structural or pathological changes (2,3). Following STandards for ReportIng Vascular changes on nEuroimaging (STRIVE) criteria (2,3), we refer to these enlarged spaces simply as PVS. In AD, increased PVS burden has been associated with clinical diagnosis and cognitive decline (6,7,12–15). However, the broader literature remains contradictory, with several studies reporting no significant relationships between PVS and AD-related biomarkers (16–18). These discrepancies are likely driven by inherent variations and limitations in PVS quantification methods (4,19–22), highlighting a critical need for robust, standardized tools capable of isolating true biological signals from imaging noise in clinical populations.

Accurate quantification and analysis of PVS requires tools that can yield reliable, granular data (e.g., whole-brain volume, precise count, and morphology). While manual delineation remains the gold standard, it is prohibitively labor-intensive for large clinical cohorts (requiring up to 20 hours per 3T scan (23)) and is subject to intra- and inter-rater variability (24,25). Visual rating scales help streamline quantification, but sacrifice the granularity required for detailed spatial analyses. Consequently, the field has rapidly shifted toward computationally derived segmentation. Recently, deep learning models have bypassed the constraints of earlier rule-based algorithms (4,19–22), offering automated feature extraction without the need for manual parameter tuning. Specifically, models built upon the self-configuring nnU-Net framework (26,27), the current state-of-the-art for medical image segmentation (26,27), have demonstrated exceptionally high performance, reporting Dice scores as high as 0.80 on internal validation datasets (23,28–31). Given this success, these automated models are increasingly poised for off-the-shelf application to clinical cohorts to investigate the relationship between PVS and neurodegeneration (4,19–22).

Despite these engineering advancements, the clinical utility of applying deep learning models to real-world AD populations remains largely unverified. The ultimate value of these tools lies in their ability to generate accurate biological inferences when deployed on external, heterogeneous clinical datasets, yet only a fraction of deep learning PVS models have been externally validated (19,20,23,29–35). Applying these models to clinical AD research introduces significant challenges. Large-scale AD studies (e.g., ADNI, A4, DIAN) utilize multi-site data, introducing scanner and acquisition heterogeneities known to alter PVS visibility (23,29,31,32). AD is also a highly heterogeneous disease characterized by concurrent structural atrophy and cerebrovascular co-pathologies, such as white matter hyperintensities (WMH), which can be misclassified as PVS. The topographical distribution of PVS also carries distinct pathological weight; for example, white matter (WM) PVS are linked to cognitive impairment, whereas basal ganglia (BG) PVS are more reflective of cerebrovascular dysfunction (5,13,36,37). If an automated segmentation model lacks regional consistency, is biased by scanner type, or fails to differentiate overlapping lesions, the downstream epidemiological relationships derived from its outputs may be spurious or misleading.

The main objective of this study was to determine whether widely available, state-of-the-art PVS segmentation models could generalize to real-world clinical cohorts, withstand neurodegenerative heterogeneity, and reliably reproduce established clinico-pathological associations. We systematically evaluated five top-performing, nnU-Net-based PVS segmentation models using a well-characterized, multi-site AD cohort (ADNI-3). Our objectives were threefold: 1) examine the segmentation performance of these models on an AD-specific dataset, alongside an assessment of broader segmentation generalizability using an external non-AD cohort; 2) determine whether model performance is robust to clinical cohort heterogeneity, specifically evaluating consistency across brain region, concurrent WMH lesion burden, diagnostic stages, scanner manufacturers, and regional PVS severity; and 3) test whether model-derived estimates of PVS burden successfully reproduce clinical associations with key AD-related biomarkers (e.g., Aβ positivity, age) found using manual labels as a proxy for downstream research validity. By systematically evaluating these models across clinically relevant parameters, we aimed to characterize their strengths and limitations and inform best practices for deploying deep learning PVS segmentation tools in AD research.

## 2. Methods

### 2.1. Study Cohorts

#### 2.1.1. Primary Cohort (ADNI-3)

Data used in the preparation of this article were obtained from the Alzheimer’s Disease Neuroimaging Initiative (ADNI) database (adni.loni.usc.edu). The ADNI was launched in 2003 as a public-private partnership, led by Principal Investigator Michael W. Weiner, MD. The primary goal of ADNI has been to test whether serial magnetic resonance imaging (MRI), positron emission tomography (PET), other biological markers, and clinical and neuropsychological assessment can be combined to measure the progression of mild cognitive impairment (MCI) and early Alzheimer’s disease (AD). Each participating site obtained institutional review board (IRB) approval, and participants provided written informed consent. Detailed inclusion, exclusion, and diagnostic criteria are published elsewhere (38).

This study utilized a curated subset of 60 participants from ADNI-3 with available co-registered 3D T1-weighted (T1w) and 3D fluid-attenuated inversion recovery (FLAIR) MRIs. Scans were acquired across over 30 different sites using Siemens (n=39), GE (n=13), and Philips (n=8) scanners. Acquisition protocols have been previously described (38). Briefly, T1w images were acquired using an accelerated 3D MPRAGE or IR-FSPGR sequence with the following parameters: repetition time (TR) = 2300ms, echo time (TE) = 2.98ms, inversion time (TI) = 900ms, flip angle 9°, field of view (FOV) = 208×240×256mm3 (with 1mm3 effective resolution). 3D FLAIR images were acquired with TR = 4800ms, TE = 119ms, TI = 1650ms, FOV = 256×256×160 mm^3^ with an effective resolution of 1.2×1×1 mm^3^.

To ensure robust evaluation across typical clinical heterogeneity, this subset was balanced across diagnostic categories (cognitively unimpaired (CU): n=20, MCI: n=20, AD: n=20). The cohort captures a wide spectrum of PVS burden across the WM and BG, varied WM hyperintensity burdens, and distribution of key AD-related biomarkers, including Aβ positivity status. Demographic and clinical characteristics are summarized in Table 1.

**Table 1:** Demographics for ADNI-3 dataset. CU, Cognitively Unimpaired; MCI, Mild Cognitive Impairment; AD, Alzheimer’s Disease; Aβ, β-amyloid; SD, standard deviation; WM, white matter; BG, basal ganglia; ICV, intracranial-volume.

|  | Total | CU | MCI | AD |
|---|---|---|---|---|
| N | 60 | 20 | 20 | 20 |
| Age (mean, std) | 75.93, 8.87 | 75.25, 5.87 | 75.60, 8.03 | 76.93, 11.99 |
| Sex (n females, %) | 27, 45.00% | 14, 70.00% | 7, 35.00% | 6, 30.00% |
| β-amyloid positivity (n Aβ+, %) | 40, 66.67% | 4, 20.00% | 17, 85.00% | 19, 95.00% |
| Hypertension (n, %) | 23, 38.33% | 5, 25.00% | 7, 35.00% | 11, 55.00% |
| WMH Volume, ICV normalized (mean, std) | 0.0068, 0.0069 | 0.0058, 0.0054 | 0.0055, 0.0049 | 0.0091, 0.0093 |
| Scanner Manufacturer (GE/Philips/Siemens; n, %) | 13 (21.67%), 8 (13.33%), 39 (65.00%) | 2 (10.00%), 2 (10.00%), 16 (80.00%) | 5 (25.00%), 4 (20.00%), 11 (55.00%) | 6 (30.00%), 2 (10.00%), 12 (60.00%) |
| PVS Severity Rating - WM (1,2,3,4; n, %) | 11 (18.33%), 20 (33.33%), 15 (25.00%), 14 (23.33%) | 3 (15.00%), 7 (35.00%), 8 (40.00%), 2 (10.00%) | 3 (15.00%), 8 (40.00%), 3 (15.00%), 6 (30.00%) | 5 (25.00%), 5 (25.00%), 4 (20.00%), 6 (30.00%) |
| PVS Severity Rating - BG (1,2,3,4; n, %) | 12 (20.00%), 24 (40.00%), 11 (18.33%), 13 (21.67%) | 4 (20.00%), 11 (55.00%), 2 (10.00%), 3 (15.00%) | 3 (15.00%), 7 (35.00%), 5 (25.00%), 5 (25.00%) | 5 (25.00%), 6 (30.00%), 4 (20.00%), 5 (25.00%) |

#### 2.1.2. External Validation Cohort (VALDO)

To assess external segmentation generalizability, a secondary, publicly available dataset was obtained from the *Where is VALDO? VAscular Lesions Detection and segmentatiOn challenge* (VALDO) challenge, run as a satellite event for the Medical Image Computing and Computer Aided Intervention (MICCAI) 2021 conference (35). This dataset consisted of 12 images with manual segmentations sourced from two distinct population-based cohorts: the Southall and Brent Revisited (SABRE) study (n=6; 3T imaging; age range 36-92 years) (39); and the Rotterdam Scan Study (RSS) (n=6; 1.5T imaging; age ≥ 45 years) (40). Both subsets provided co-registered and skull-stripped T1w, T2w, and FLAIR images. Ground-truth manual segmentations were provided for randomly selected 5mm slabs (SABRE) or specific slices in the centrum semiovale and BG (RSS) (example regional selections are shown in Supplementary Figure S1). Since the SABRE images were segmented by two raters, we combined their masks into a single reference mask via a logical OR such that a voxel was included in the reference mask if either rater labeled it.

### 2.2. Manual Segmentation of Reference Set & Image Preprocessing

Manual PVS segmentation of the whole brain from the ADNI-3 cohort was performed by a trained rater (S.T.) under the guidance of an experienced neurologist (K.Y.) using ITK-SNAP (version 4.2.0), adhering to STRIVE criteria (2,3,24). To avoid the misclassification of overlapping pathologies, particularly WMH, co-registered T1w and FLAIR images were evaluated simultaneously. Inter-rater reliability was assessed via independent whole-brain manual labeling by a second trained rater (E.P.) on a subset of 5 images, which was further verified by K.Y. Intra-rater reliability was assessed by S.T. re-evaluating the same subset after a one-year interval. For both, intraclass correlation coefficients (ICC) were calculated to determine concordance, where intra-rater used two-way mixed effects for single rater and inter-rater used two-way random effects (41).

To evaluate region-specific performance, automated segmentations were masked to WM (using the *wmparc.mgz* mask obtained from FreeSurfer version 6.0.0) and BG (using the *aparc+aseg.mgz* mask), inclusive of the caudate, putamen, globus pallidus, nucleus accumbens, and thalamus). To determine if models erroneously segmented WMH as PVS, WMH masks were generated from FLAIR images using the Lesion Prediction Algorithm (LPA) from the LST toolbox in SPM12 (LST toolbox version 3.0.0 for SPM (42,43), Chapter 6.1 (43)) (example is shown in Supplementary Figure S1).

### 2.3. Deep Learning Models

We utilized the nnU-Net (“no-new U-Net”) framework as the base architecture due to its state-of-the-art performance in biomedical image segmentation benchmarks (26,27) as well as PVS segmentation tasks (6,23,28–32). The nnU-Net is a U-Net based segmentation method that employs on-the-fly data augmentation and self-configuration of preprocessing, training and postprocessing parameters to optimally segment a wide variety of medical images (26,27,44).

For this study, open-source PVS segmentation models were included if they: 1) performed 3D segmentations encompassing both the WM and BG; 2) used an nnU-Net based framework; and 3) did not require more than either T1w or T1w and FLAIR images. A comprehensive list of reviewed models and inclusion criteria is provided in Supplementary Table S1.

Five models were evaluated: nnU-Net T1 and nnU-Net T1+FLAIR (28), mcPVS-Net (23), MedNet-PVS (31), and ADNI-PVS-Net (6) (Table 2), a newer model optimized for AD populations. PINGU (29) was excluded to prevent redundancy, as it shares training data with MedNet-PVS but uses an older nnU-Net iteration (nnunetv1). All models can be accessed here: https://github.com/PVS-segmentation-repository.

**Table 2:**
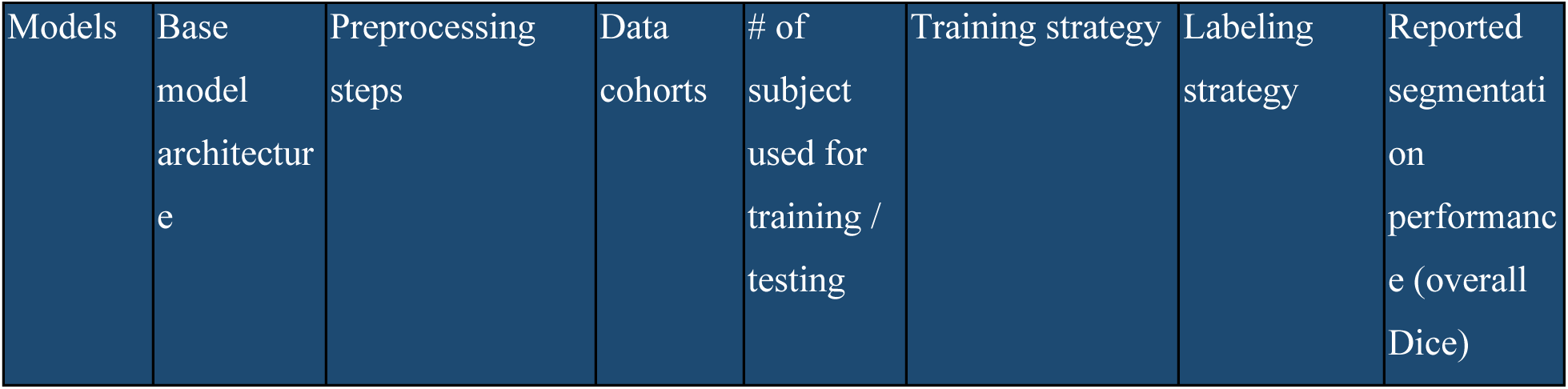
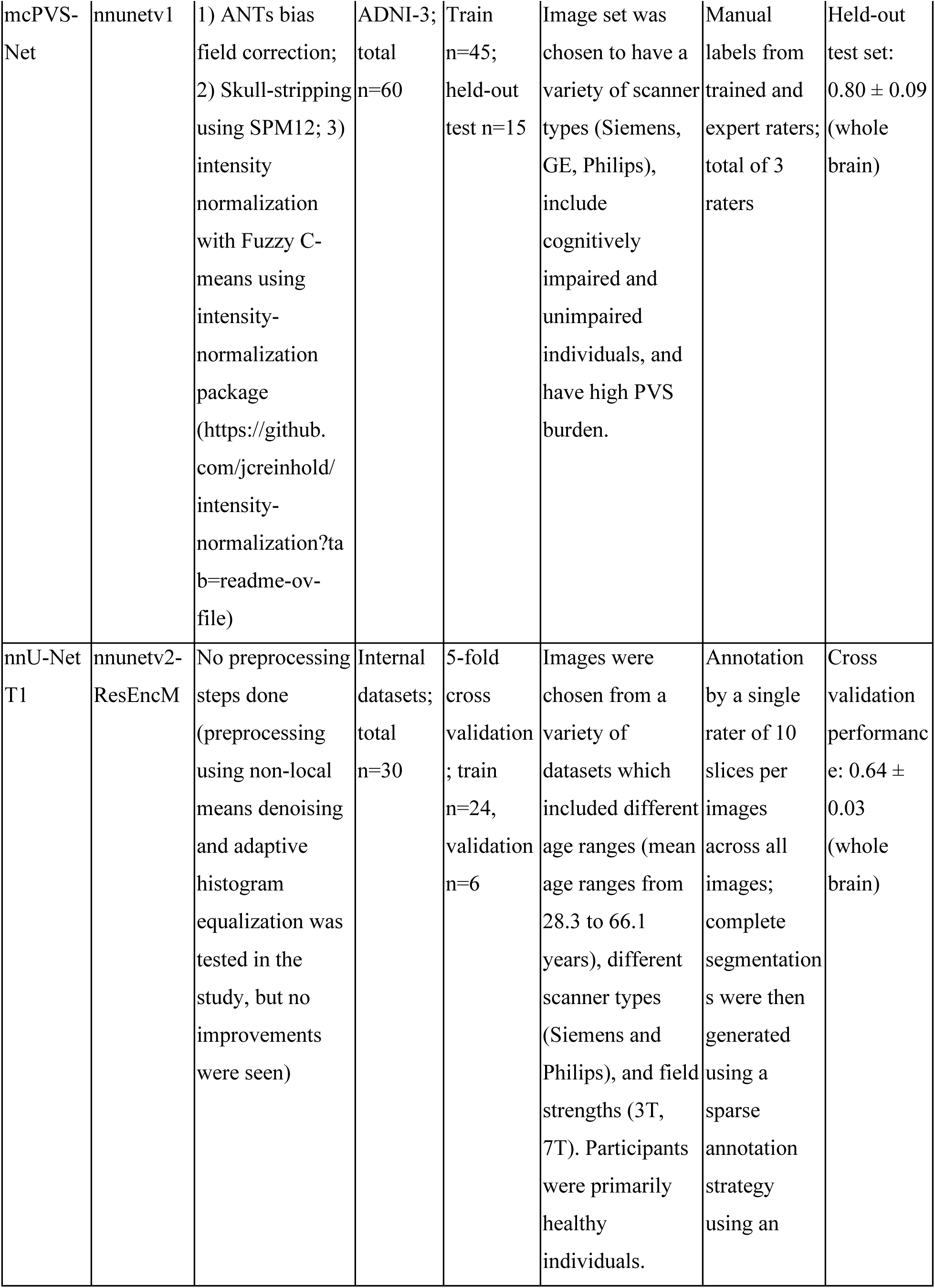
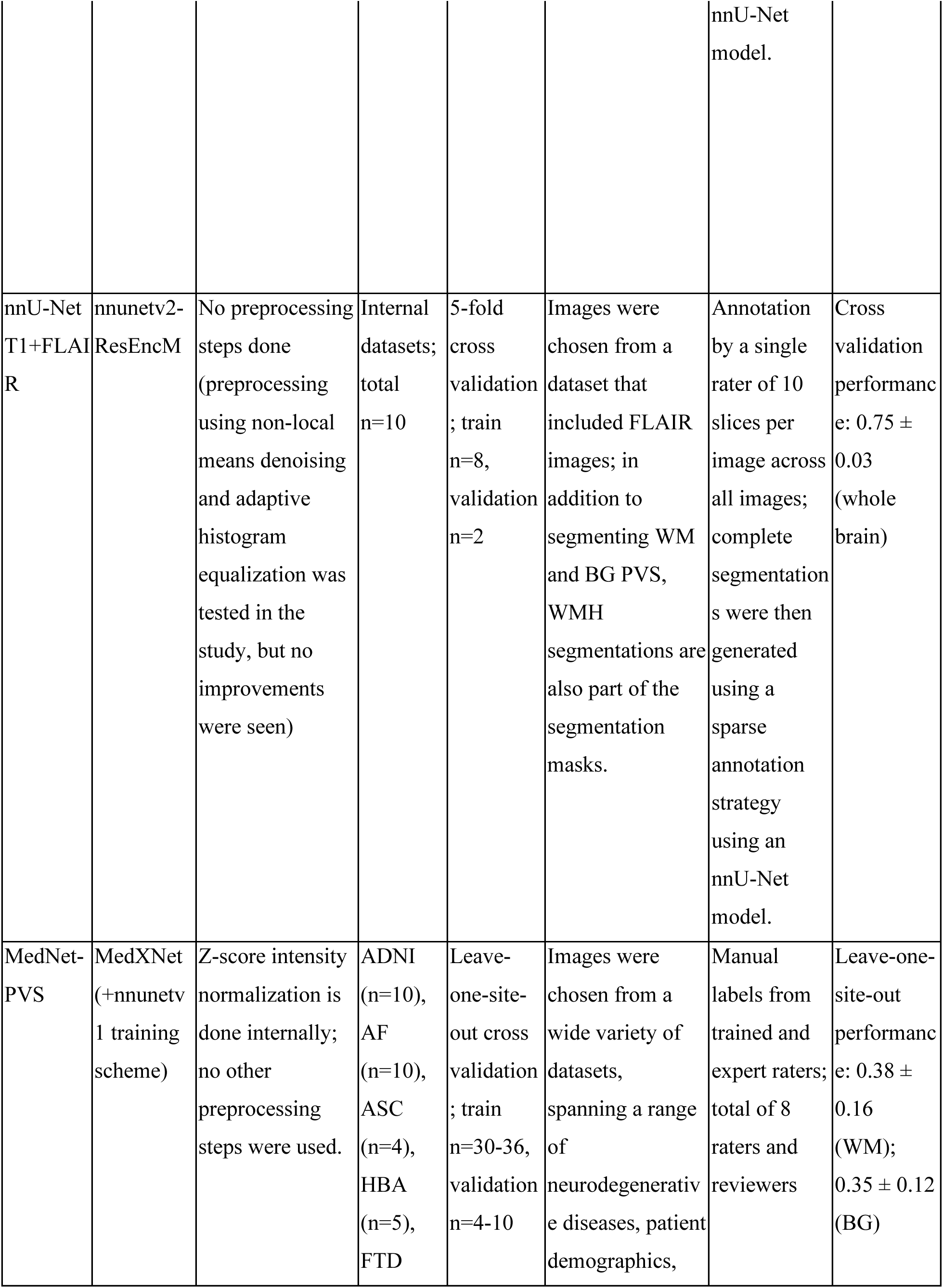
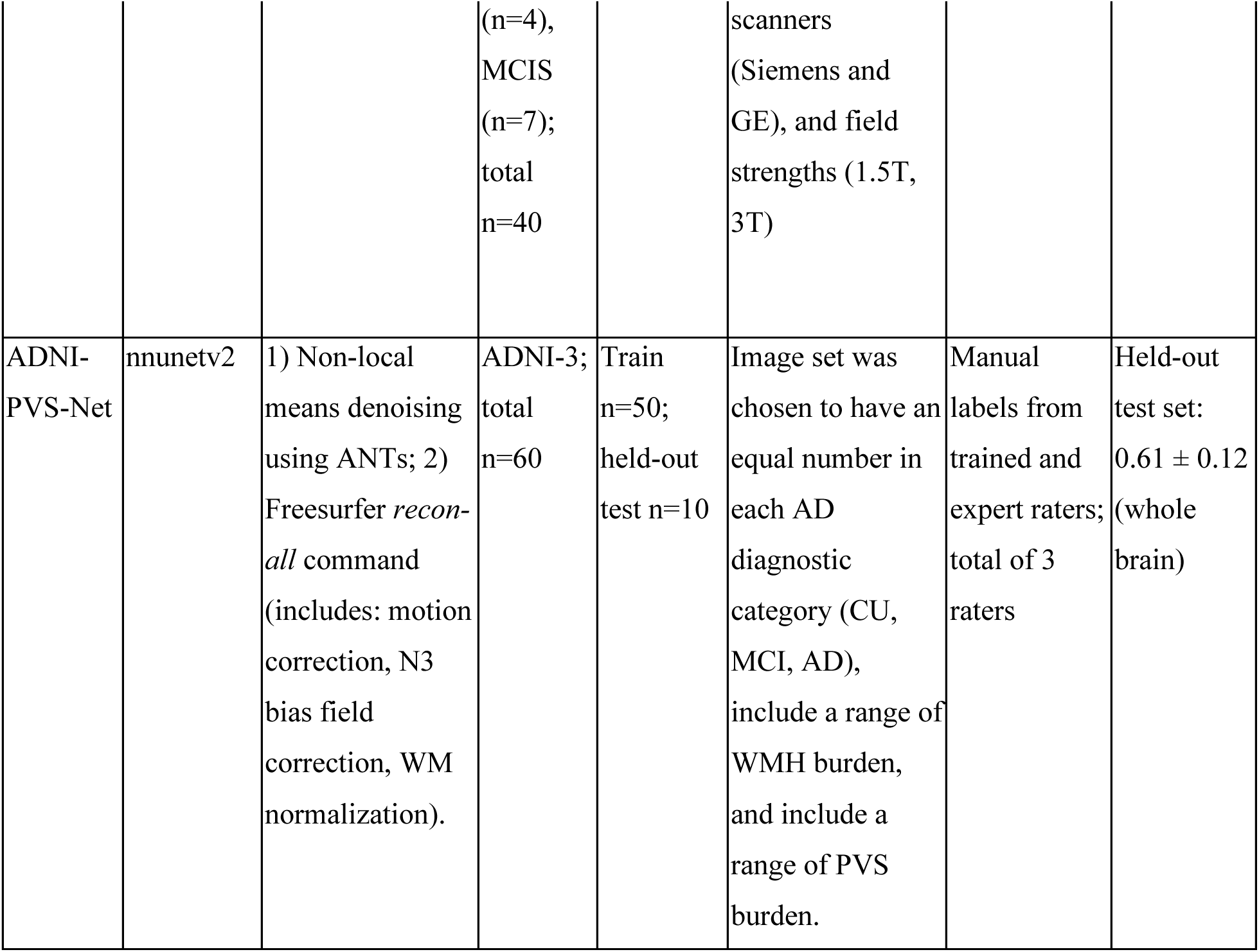
nnU-Net Model Development Descriptions. Descriptions of each model, their base architecture, preprocessing steps, dataset used, training and test splits, training strategy, labeling strategy, and reported internal performance.

Since ADNI-PVS-Net was developed natively on the 60 ADNI-3 scans included in this study, predicting on its own training data would artificially inflate clinical performance. To prevent data leakage and provide an unbiased evaluation for downstream clinical comparisons, we implemented a strict 6-fold cross-validation scheme. The model was trained six separate times on six different subsets of 50 subjects, generating unseen, out-of-fold inference for 10 held-out subjects each time; these were then combined to form the full 60 subject set. All reported results for ADNI-PVS-Net on the ADNI cohort reflect these unbiased out-of-fold predictions. For VALDO, a separate ADNI-PVS-Net model trained once on all 60 subjects was used; this also reflects the publicly available version of this model.

Raw T1w and co-registered FLAIR images (FSL FLIRT version 6.0.7.13, 6-degrees of freedom) served as standard inputs. Subsequent preprocessing steps (e.g., N3/N4 bias field correction, skull-stripping, intensity normalization) were rigorously replicated exactly as prescribed by the respective developers of each model (Table 2).

### 2.4 Performance Evaluation Framework

#### 2.4.1 Baseline segmentation metrics (Aim 1)

Because standard whole-brain spatial overlap may not fully capture the clinical utility of PVS delineation (e.g., counting individual lesions vs. volumetric precision) (45), performance was evaluated using four distinct Dice similarity coefficient (DSC) metrics (illustrated in Supplementary Figure S2):

Overall Dice: The standard voxel-wise spatial overlap between predicted and ground-truth masks across the entire region of interest.

Object-level Dice (Instance Detection Rate): Evaluates the model’s ability to detect individual PVS lesions regardless of precise segmentation boundaries. A true positive is defined as any spatial overlap (>0 voxels) between a predicted PVS cluster and a ground-truth PVS instance.

Voxel-wise Dice: Assesses precise boundary delineation only for successfully detected lesions. Standard Dice is calculated exclusively within the union of predicted PVS that successfully intersect with a ground-truth PVS, ignoring false-positive and false-negative instances.

Lesion-wise Dice: Evaluates performance consistency across varying PVS sizes. Standard Dice is calculated independently for every individual true-positive lesion and then averaged, granting equal weight to small and large PVS.

For all metrics, equation (1) was used to calculate the Dice score (where TP = true positive, FP = false positive, and FN = false negative).

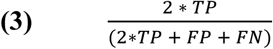

#### 2.4.2 Robustness to cohort heterogeneity (Aim 2)

To assess model robustness to clinical heterogeneity, metrics were stratified across diagnostic stage (CU, MCI, AD), scanner manufacturer (Siemens, Philips, GE), and brain region (WM vs BG). To evaluate consistency across varying PVS burden, we adapted established semi-quantitative visual rating scales for T1w images (24,25). The single axial slice above the anterior commissure containing the maximum bihemispheric PVS count was identified independently for the WM and BG. These maximum slice counts correlated strongly with whole-brain PVS burden (Spearman r=0.93, p<0.001). The distribution was then divided into quartiles to define a quantitative 4-tier severity rating. Model-predicted masks underwent the identical procedure to define model-derived severity ratings.

Within the subset of cases exhibiting PVS within WMH (n=33, quantified as PVS overlapping with WMH masks (6)), models were evaluated on the false discovery rate (the proportion of predicted PVS within the WMH mask that were actually WMH) and the false positive rate normalized by total WMH volume (the extent of PVS missegmentation given a WMH volume).

#### 2.4.3. Association with biomarkers (Aim 3)

To determine whether automated models could successfully reproduce clinical associations found via manually labeled images, relationships between PVS burden and clinical biomarkers (age, sex, Aβ-positivity, history of hypertension, and WMH volume) were modeled. For PVS and WMH volume, we used intracranial volume-normalized volumes. Due to their overdispersed, right-skewed distributions, PVS counts were analyzed using Negative Binomial regression. Continuous PVS volumes were analyzed using generalized linear models (GLMs) with a Gamma distribution to accommodate strictly positive, right-skewed data. Residual normality was verified via visual inspection of residual plots and the quantile residual Shapiro Wilks test. In this study, we report exponentiated coefficients (β_exp_) for both Negative Binomial and Gamma GLMs, which can be interpreted as the multiplicative change (rate ratio for counts, fold change for volumes) in the expected outcome for one unit change in a given covariate, or a change from the reference group for categorical variables. All p-values within biomarker regression families and pairwise model comparisons were adjusted for multiple comparisons using the Benjamini-Hochberg False Discovery Rate (FDR) correction.

### 2.5. Statistical Analysis

To evaluate the rank preservation and absolute agreement of PVS counts and volumes between predicted and ground-truth masks, we calculated Spearman rank correlation coefficients (ρ) and Lin’s concordance correlation coefficients (CCC). Statistically significant differences in segmentation scores between pairs of models were assessed using Wilcoxon signed-rank tests, while differences between clinical groups (i.e. diagnostic, scanner, PVS severity) were assessed using Mann Whitney-U tests. Agreement between model-derived and ground-truth categorical PVS severity ratings was assessed using weighted Cohen’s kappa (*κ*) and percent agreement. All statistical analyses were evaluated at a significance threshold of ɑ=0.05. All statistical computation was done using Python version 3.12; statistical modeling was done using the *statsmodels* package version 0.15.0 (47).

## 3. Results

### 3.1 Baseline Segmentation Performance and External Generalizability

#### 3.1.1. Internal Validation on ADNI-3

To determine the validity of ground truth rating methods, we calculated ICC to determine intra- and inter-rater reliability. Intra-rater reliability was 0.83 for PVS count and 0.85 for PVS volume, while inter-rater reliability was 0.75 for PVS count and 0.96 for PVS volume, indicating good to excellent reliability of rating methods (41).

Example segmentations by each model compared to the original image and reference segmentations are shown in Figure 1; numerical results are summarized in Supplementary Table S2. Segmentation performance across the primary ADNI-3 cohort varied substantially depending on the model and the specific metric evaluated (Figure 3; Supplementary Table S2). Unless otherwise noted, all pairwise comparisons between models were statistically significant (p<0.05; full pairwise results in Supplementary Table S2).

**Figure 1:**
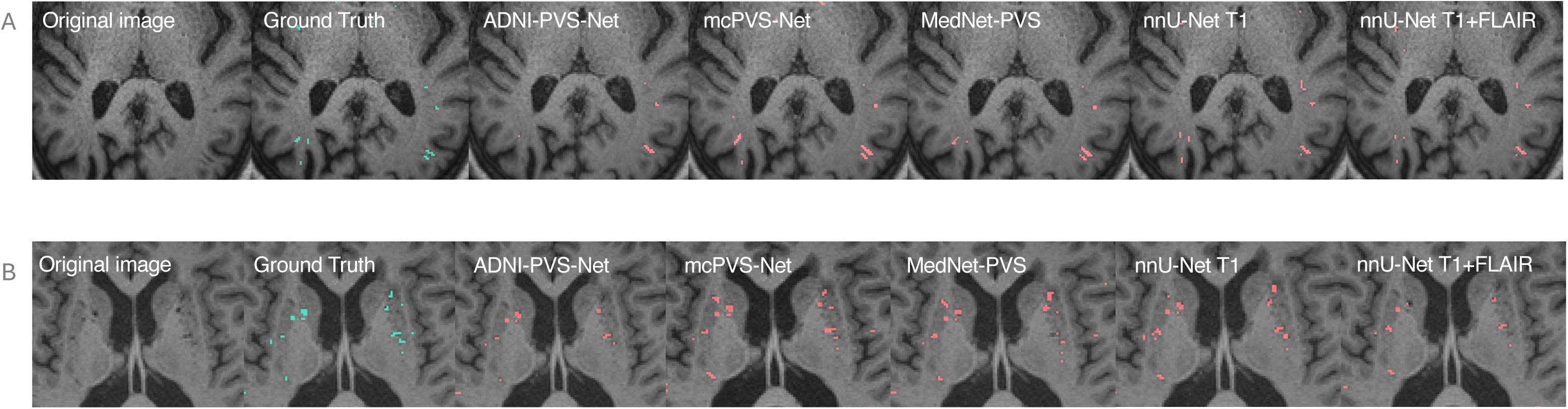
Example segmentations from models within the ADNI-3 dataset. (A) Shows each model’s segmentation in white matter compared to ground truth and original image, while (B) shows each model’s segmentation in basal ganglia.

Evaluated strictly via unbiased out-of-fold predictions to prevent data leakage, ADNI-PVS-Net achieved the highest overall spatial overlap (overall Dice: 0.58 ± 0.14), instance detection rate (object-level Dice: 0.66 ± 0.14), and boundary precision for detected lesions (voxel-wise Dice: 0.73 ± 0.12), outperforming every other model on all three metrics. The next-best model for spatial overlap was MedNet-PVS (overall Dice 0.29 ± 0.15), which differed significantly from ADNI-PVS-Net and mcPVS-Net only. However, evaluation of lesion-wise Dice, which weights small and large PVS equally, revealed a different hierarchy: MedNet-PVS (0.79 ± 0.16) and mcPVS-Net (0.76 ± 0.19) did not differ significantly from each other but outperformed all other models, while ADNI-PVS-Net (0.53 ± 0.18) underperformed relative to every model except nnU-Net T1. This suggests a performance trade-off where ADNI-PVS-Net excels at capturing total whole-brain PVS burden, whereas MedNet-PVS and mcPVS-Net delineate the boundaries of individual (especially smaller) PVS with higher uniform precision despite missing a larger total number of lesions.

All models showed moderate-to-strong, statistically significant Spearman rank correlations with ground-truth PVS counts (ρ = 0.54-0.92) and volumes (ρ = 0.55-0.92, all p < 0.001; Supplementary Table S2). However, absolute agreement (Lin’s CCC) revealed wider disparities. ADNI-PVS-Net showed the strongest absolute agreement (count CCC = 0.86, volume CCC = 0.61), whereas mcPVS-Net and MedNet-PVS showed relatively weak absolute agreement (count CCC = 0.28, volume CCC = 0.30) despite highly preserved rank ordering, suggesting systematic under- or over-estimation of the total burden.

#### 3.1.2 External Generalizability on VALDO

To assess out-of-distribution generalizability, models were evaluated on the independent VALDO dataset (1.5T RSS and 3T SABRE subsets); example segmentations and selected regions are shown in Figure 2; numerical results are summarized in Supplementary Table S3; graphical results are shown in Figure 3. Performance degraded universally across all models compared to ADNI-3. Notably, ADNI-PVS-Net, despite its effectiveness on the ADNI-3 cohort, exhibited the most severe generalizability drop on both VALDO subsets (overall Dice: 0.11 ± 0.07 on RSS; 0.04 ± 0.03 on SABRE).

**Figure 2:**
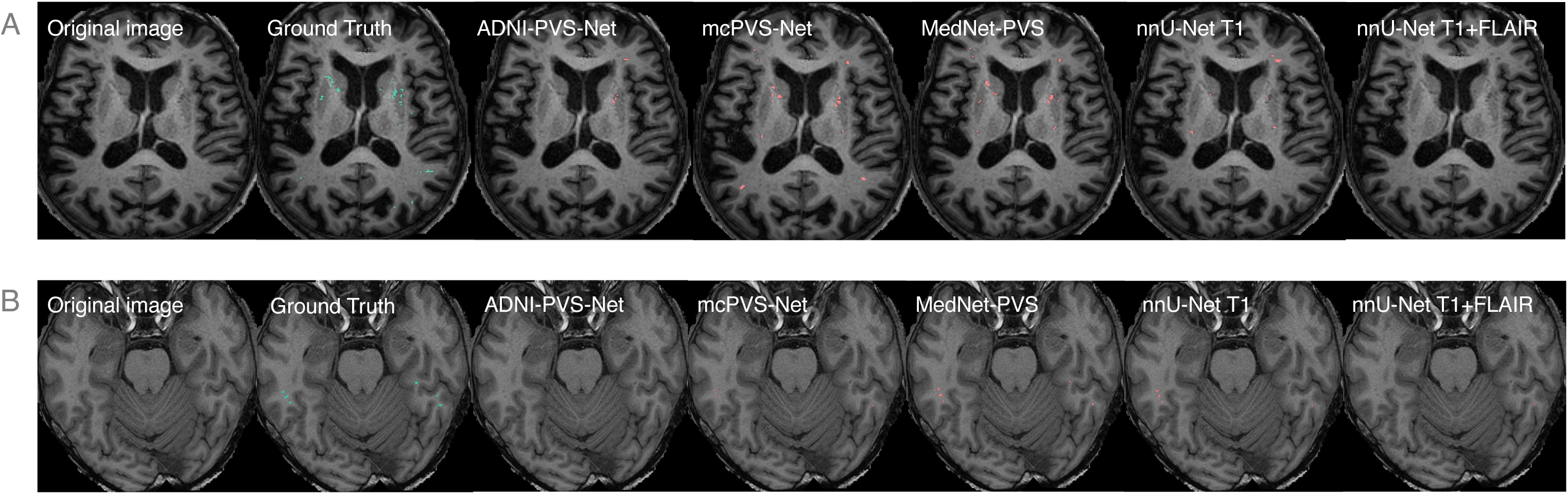
Example segmentations from models within the VALDO dataset. (A) Shows model’s predictions on the SABRE dataset, while (B) shows predictions on the RSS dataset.

Conversely, mcPVS-Net and MedNet-PVS demonstrated higher relative resilience, maintaining or slightly exceeding their ADNI-3 baseline performance on the 1.5T RSS subset, and emerging as the top performers on the external data (overall Dice, mcPVS-Net: 0.27 ± 0.14; MedNet-PVS: 0.30 ± 0.09). mcPVS-Net differed significantly only from ADNI-PVS-Net and nnU-Net T1+FLAIR (p<0.05), while MedNet-PVS differed significantly from all models except mcPVS-Net (p<0.05). On the other hand, models performed relatively consistently when evaluated by voxel-wise Dice, ranging from 0.39-0.55, with no significant pairwise differences except between mcPVS-Net and nnU-Net T1+FLAIR (p<0.05). Correlations between model-derived and ground-truth metrics were largely weak and non-significant on VALDO, apart from mcPVS-Net and MedNet-PVS volumetric estimates, which reached significance on the RSS subset (both ρ = 0.83, p = 0.04; Supplementary Table S3).

### 3.2 Robustness to Cohort Heterogeneity

#### 3.2.1 Regional performance and WMH discrimination

Overall Dice scores dropped by 0.05 to 0.20 points in the BG compared to the WM across all models except mcPVS-Net, which retained stable performance across both regions (Figure 3; Supplementary Table S4). ADNI-PVS-Net maintained the highest overall and object-level Dice in both regions, and overall model ranking and comparisons were consistent with overall performance (Figure 3; Supplementary Table S4). Rank correlations remained strong in the WM but weakened considerably in the BG; notably, nnU-Net T1+FLAIR entirely failed to preserve rank correlations for BG counts or volumes (both p > 0.90).

**Figure 3:**
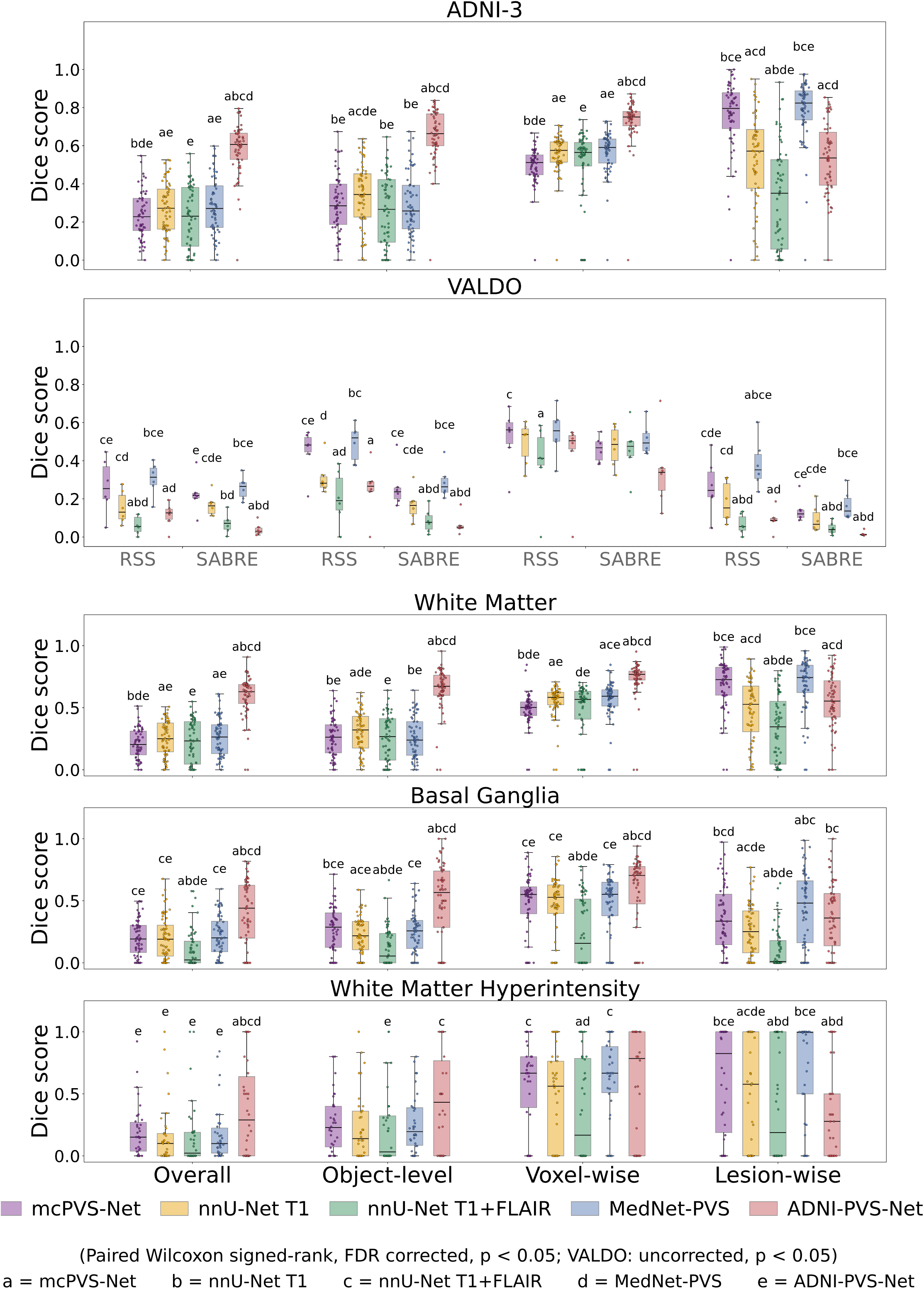
Grouped box plots comparing model performance across the internal (ADNI-3) and external (VALDO) test datasets, and across brain region (white matter, basal ganglia, and white matter hyperintensity), for four Dice-based metrics: overall, object-level, voxel-wise, and lesion-wise Dice. Letters above each box indicate the other models it differs significantly from in pairwise comparison (paired Wilcoxon signed-rank test, FDR-corrected for ADNI-3, p < 0.05; see legend for the letter-to-model key). ADNI-PVS-Net scored significantly higher than the other four models on overall, object-level, and voxel-wise Dice across ADNI-3 and all regional comparisons, followed by mcPVS-Net and MedNet-PVS, while the latter two models had higher scores on lesion-wise Dice. Performance dropped for most models on the external VALDO dataset, except for mcPVS-Net and MedNet-PVS, which performed similarly to their ADNI-3 results. Notably, ADNI-PVS-Net’s performance dropped most sharply on VALDO (overall Dice: ADNI-3 = 0.58 ± 0.14; VALDO-RSS = 0.11 ± 0.07; VALDO-SABRE = 0.04 ± 0.03).

Discrimination of true PVS from spatially adjacent WMH proved highly challenging for T1-only models (Figure 3; Supplementary Table S5). MedNet-PVS exhibited the highest false discovery rate (0.81 ± 0.20) and misclassified the largest volume of true WMH as PVS (over 32% of WMH volume). Conversely, models trained utilizing multi-modal inputs, specifically ADNI-PVS-Net and nnU-Net T1+FLAIR, successfully minimized false positives within ground-truth WMH boundaries, yielding normalized false positive rates of just 1.04% and 11.73% respectively (Supplementary Table S5).

#### 3.2.2. Robustness to disease stage and scanner manufacturer

All five models demonstrated robust consistency across diagnostic stages (CU, MCI, AD), with no significant differences in segmentation metrics across clinical groups (all p ≥ 0.17, Figure 4, Supplementary Table S6). However, robustness across scanner manufacturers varied significantly (Figure 4; Supplementary Table S7). nnU-Net T1+FLAIR showed the most pronounced scanner-dependent degradation, with overall Dice dropping from 0.30 ± 0.13 on Siemens scanners to 0.03 ± 0.05 on Philips scanners (p < 0.001). MedNet-PVS and nnU-Net T1 also showed statistically significant scanner effects across multiple metrics (p ≤ 0.02). mcPVS-Net and ADNI-PVS-Net were the most scanner-agnostic, exhibiting stable overall, object-level and voxel-wise performance across all hardware manufacturers.

**Figure 4:**
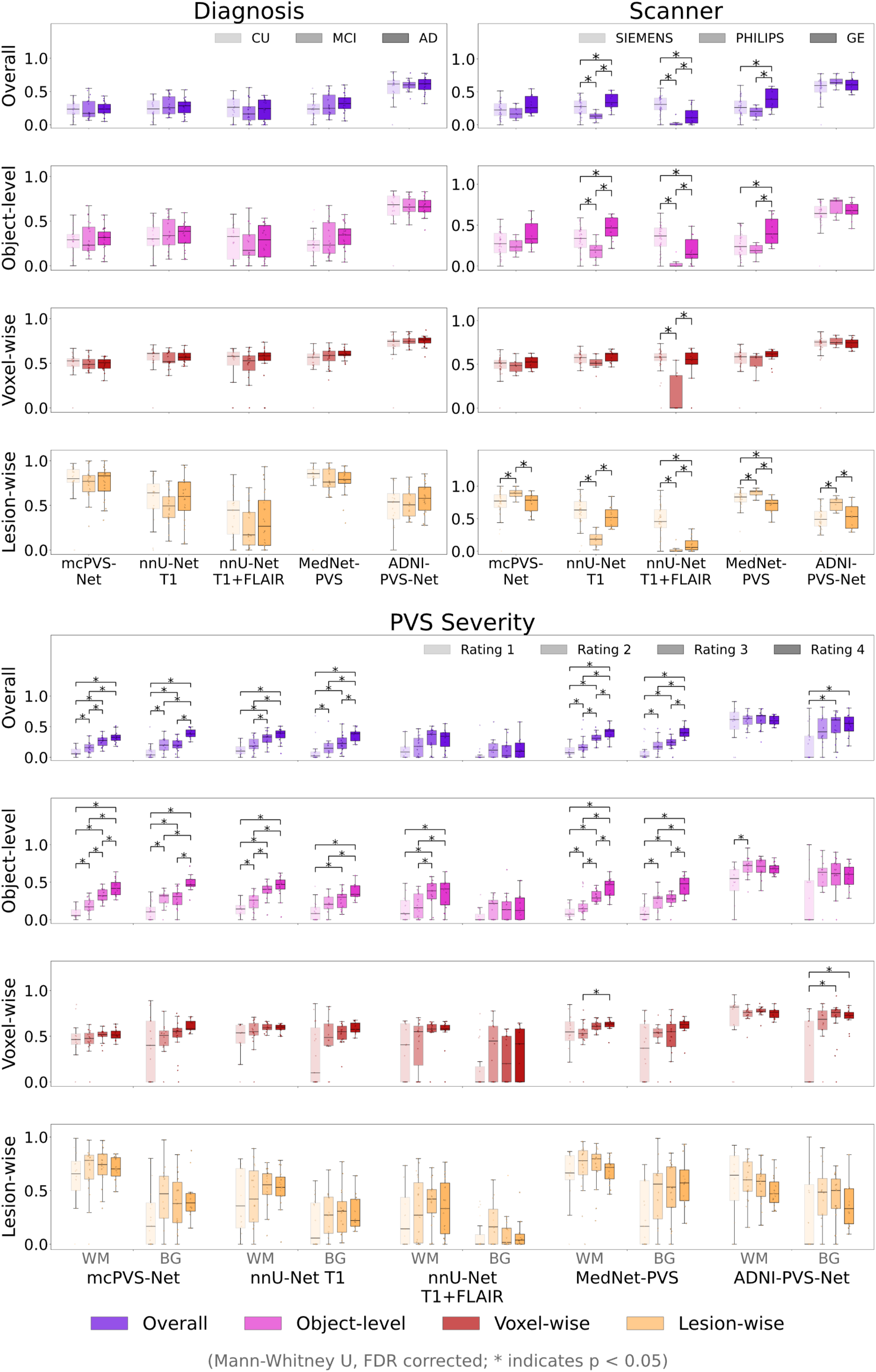
Grouped box plots show model robustness across diagnostic, scanner, and PVS severity categories. Each panel column corresponds to one clinical category (diagnosis, scanner, or PVS severity), and each row corresponds to one Dice-based performance metric (overall, object-level, voxel-wise, lesion-wise). Within each panel, the group of boxes for a given model shows that model’s performance across the category’s subgroups (e.g., CU/MCI/AD for diagnosis), indicated by shading from lightest to darkest. Brackets connect pairs of subgroups with a significant difference in Dice score within that model (Mann-Whitney U test, FDR-corrected, p < 0.05), with a star marking each significant pair. All models were robust to diagnostic groups and mostly robust across scanner manufacturers, though nnU-Net T1 and nnU-Net T1+FLAIR showed significant differences across scanners. mcPVS-Net and ADNI-PVS-Net remained largely robust except in lesion-wise Dice. Across PVS severity, ADNI-PVS-Net was most robust overall, though it showed some sensitivity in object-level Dice within white matter; the other four models varied significantly by severity in overall and object-level Dice. All models were largely robust to severity on voxel-wise and lesion-wise Dice.

#### 3.2.3. Consistency across PVS severity

Model efficacy was heavily influenced by the underlying PVS burden (Figure 4; Supplementary Tables S8A, S9A). For most models, overall and object-level Dice increased significantly as visual PVS severity increased from rating 1 (lowest burden) to rating 4 (highest burden) (p < 0.001; Supplementary Tables S8A, S9A). This indicates that most standard architectures struggle to accurately segment sparse PVS in relatively healthy subjects. ADNI-PVS-Net was the notable exception, maintaining statistically stable overall, voxel-wise, and lesion-wise Dice across all severity quartiles in the WM, though its object-level detection still scaled with PVS burden.

When converting predicted outputs into clinical severity quartiles, ADNI-PVS-Net showed the strongest absolute agreement with ground-truth manual ratings (WM: weighted κ = 0.85, 68.33% agreement; BG: κ = 0.78, 66.67% agreement; Supplementary Table S8B, S9B). The remaining models showed only slight-to-moderate clinical agreement (WM range: weighted κ = 0.12-47, 28-46% agreement; BG range: weighted κ = 0.11-43, 28-46% agreement; Supplementary Table S8B, S9B).

### 3.3 Association with biomarkers

To determine downstream research validity, we assessed whether predicted PVS burdens could reliably replicate ground-truth clinical associations (Figure 5; numerical results are summarized in Supplementary Table S10A-C). In the manual ground-truth data, PVS counts were significantly associated with Aβ-positivity (Aβ+ vs. Aβ−) globally (β_exp_ = 2.46, CI=[1.44, 4.22], p = 0.01), in WM (β_exp_ = 2.48, CI=[1.36, 4.53], p = 0.02), and in the BG (β_exp_ = 2.25, CI=[1.25, 4.05], p = 0.04), as well as PVS volumes (global: β_exp_ = 2.72, CI=[1.45, 5.14], p = 0.01; WM: β_exp_ = 2.60, CI=[1.33, 5.07], p = 0.02). No significant associations were found with WMH volume, sex, or hypertension (all p ≥ 0.06).

**Figure 5:**
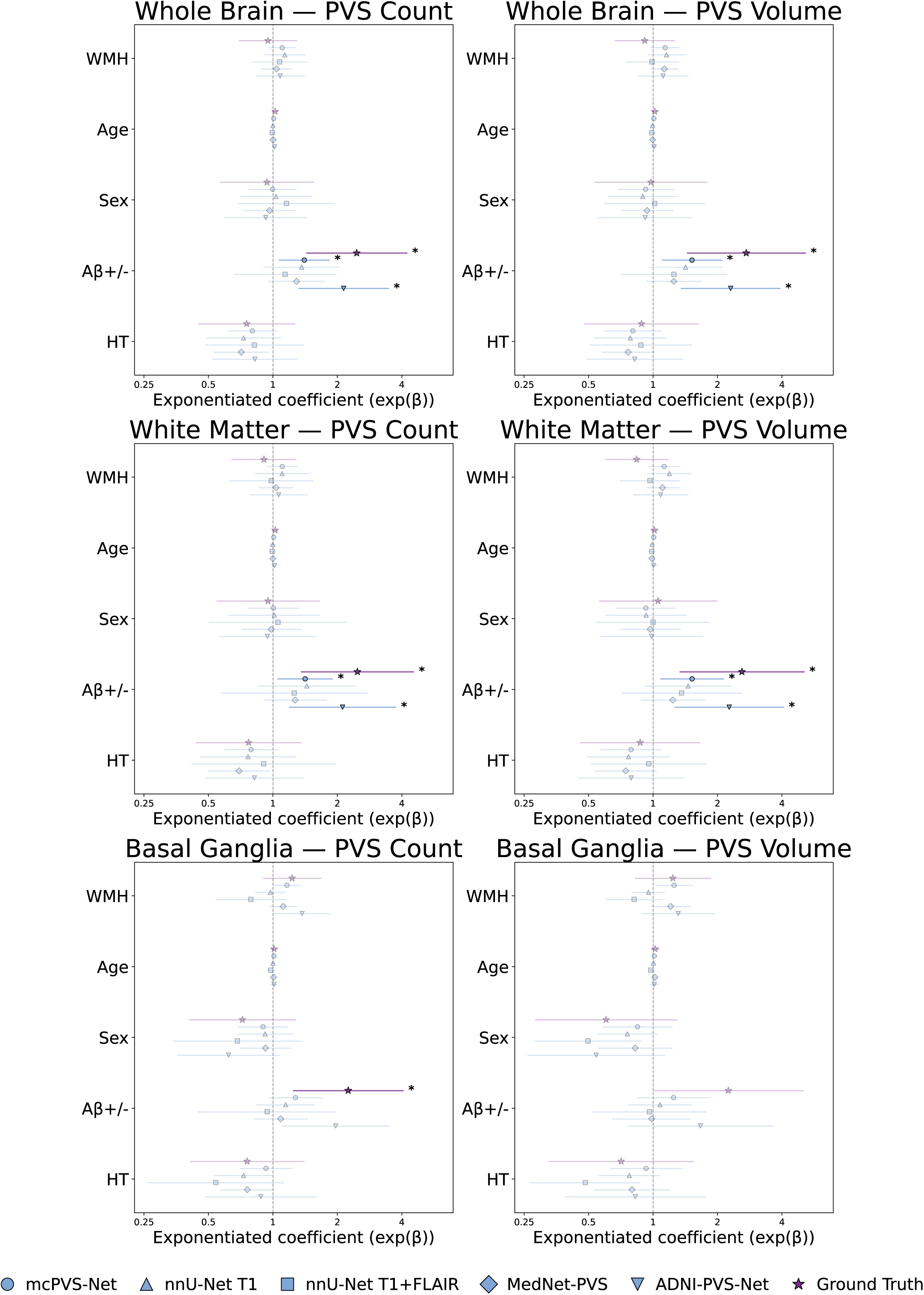
Forest plots show the association between clinical/demographic covariates (WMH volume, age, sex, Aβ positivity, hypertension) and PVS count or volume across models. Exponentiated coefficients (points) and 95% confidence intervals (lines) represent the multiplicative change in expected PVS count or volume per covariate (values > 1 indicate an increase; Aβ status uses Aβ− as the reference). The purple star at the top of each row shows the ground truth estimate. Faded points/lines are non-significant, and opaque points with an asterisk are significant (FDR-corrected, p < 0.05). Only models trained on AD-specific data reproduced the ground truth’s significant relationships, suggesting domain-specialized training may be needed to capture disease effects accurately.

Automated models reliably mirrored the absence of associations with WMH, sex, and hypertension, producing no false-positive clinical findings. However, their sensitivity to detect true-positive relationships was heavily limited (Figure 5; Supplementary Tables S10A-C). Notably, only the two models explicitly trained on AD cohorts successfully replicated the significant relationship between Aβ-positivity and PVS burden: mcPVS-Net (count global: β_exp_ = 1.40, CI=[1.07, 1.83], p = 0.03; count WM: β_exp_ = 1.41, CI=[1.06, 1.90], p = 0.04; volume global: β_exp_ = 1.52, CI=[1.11, 2.09], p = 0.02; volume WM: β_exp_ = 1.52, CI=[1.09, 2.13], p = 0.03) and ADNI-PVS-Net (count global: β_exp_ = 2.14, CI=[1.32, 3.46], p = 0.01; count WM: β_exp_ = 2.12, CI=[1.20, 3.74], p = 0.03; volume global: β_exp_ = 2.30, CI=[1.35, 3.92], p = 0.01; volume WM: β_exp_ = 2.27, CI=[1.27, 4.06], p = 0.02; Supplementary Table S10A, S10B), with ADNI-PVS-Net most closely recovering the effect size. nnU-Net T1, nnU-Net T1+FLAIR, and MedNet-PVS completely failed to detect this primary relationship (all p ≥ 0.10). No models were able to replicate the significant relationship with Aβ-positivity in the BG (all p > 0.05, Supplementary Table S10C).

## 4. Discussion

With the exponential rise in the application of open-source, deep learning-based segmentation models to investigate the role of PVS in neurodegenerative diseases (4), it is crucial to ensure these tools are robust when considering the heterogeneity of clinical cohorts. This study is the first to systematically evaluate state-of-the-art nnU-Net PVS segmentation models across clinically relevant dimensions in an Alzheimer’s-related cohort. Overall, we observed a trade-off between domain-specific optimization and out-of-distribution generalizability. Models trained explicitly on AD populations (ADNI-PVS-Net, mcPVS-Net) successfully replicated ground-truth epidemiological associations on the ADNI cohort, but struggled on external datasets. Conversely, models trained on highly diverse, non-AD cohorts (MedNet-PVS, nnU-Net T1) were more resilient to external domain shifts but failed to capture critical disease-specific biomarker relationships within the AD population. These findings show that strong internal performance metrics do not automatically translate to downstream clinical utility.

### 4.1. Internal Performance vs. External Generalizability

On the ADNI-3 data, the ADNI-PVS-Net model substantially surpassed other models across almost all metrics; however, its performance dropped considerably when evaluated on the external VALDO dataset. In contrast, mcPVS-Net and MedNet-PVS achieved lower but consistent scores across both ADNI-3 and VALDO datasets. These results illustrate a domain shift failure: ADNI-PVS-Net acts as a domain specialist model that is highly tuned to the specific imaging characteristics of the ADNI dataset, while MedNet-PVS and mcPVS-Net function as “generalists” that exhibit modest but stable performance on out-of-distribution data. Although all evaluated models employed strategies intended to improve out-of-distribution generalizability, such as training on diverse sites, scanners, diseases, ages, and PVS burden, these results underscore the persistent vulnerability of deep learning models to domain shifts, highlighting the necessity of external validation before deploying publicly available models.

Examining specific segmentation metrics clarifies where these models succeeded and faltered. ADNI-PVS-Net excelled at capturing the total whole-brain PVS burden (high overall and object-level Dice) but ranked fourth in lesion-wise Dice, suggesting a trade-off between capturing the total volume and precisely delineating individual boundaries. MedNet-PVS showed the opposite pattern: it delineated the individual PVS instances it found with high precision (highest lesion-wise Dice; Figure 3, Supplementary Table S2) but missed a substantial number of lesions overall (lower object-level Dice; Figure 3, Supplementary Table S2). Despite being trained on a younger, healthy cohort, the nnU-Net T1 model achieved competitive performance, suggesting that its training diversity and residual encoder architecture meaningfully contributed to its generalizability. Conversely, nnU-Net T1+FLAIR yielded the lowest overall spatial overlap, consistent with its original developmental validation which noted a slight performance reduction when adding the FLAIR modality.

The drop in performance on the external VALDO dataset further illustrates domain shift as a failure point for these models. Both nnU-Net T1+FLAIR and ADNI-PVS-Net, which used FLAIR in addition to T1 at inference, scored the lowest, potentially reflecting a disadvantage of including FLAIR during training. mcPVS-Net and MedNet-PVS, by contrast, were trained on datasets with higher PVS burden and greater variety in manual delineation, likely contributing to their more stable performance. Both models also performed similarly across the ADNI and RSS datasets despite the difference in field strength, suggesting some robustness to field strength changes. This robustness did not extend to the SABRE dataset, however: all models performed worse on SABRE than on RSS despite SABRE matching the models’ training field strength, suggesting other image characteristics not addressed by each model’s preprocessing pipeline. Nevertheless, these results should be interpreted with some caution, as overall sample size was small, assessment was limited to specific slabs or slices rather than the whole brain, and images were pre-skull-stripped, preventing exact replication of preprocessing for the three models (ADNI-PVS-Net, nnU-Net T1, and nnU-Net T1+FLAIR) trained exclusively on non-skull-stripped images.

### 4.2. Robustness to Clinical Heterogeneity

Because AD patients exhibit varied co-pathology, PVS burden often concentrates in distinct brain regions associated with different mechanisms (e.g., WM-PVS with cognitive impairment; BG-PVS with cerebrovascular dysfunction). Almost all models performed worse in the basal ganglia than in the white matter, likely due to the region’s smaller anatomical search space and lower tissue contrast.

Distinguishing PVS from WMH remains a fundamental challenge for automated segmentation methods (3,4). Because WMH is ubiquitous in AD, avoiding false positives is critical. Models trained strictly on T1-weighted images struggled profoundly; MedNet-PVS misclassified a substantial portion of true WMH volume as PVS despite being trained on a clinical cohort. Conversely, multi-modal architectures (ADNI-PVS-Net and nnU-Net T1+FLAIR) successfully utilized FLAIR data to minimize these false positives. Based on these results, multi-channel models are recommended for AD cohorts to prevent overlapping vascular lesions from artificially inflating PVS estimates.

Although models had robust performance across diagnostic categories, performance varied across hardware and PVS severity. While mcPVS-Net and ADNI-PVS-Net were relatively scanner-agnostic, nnU-Net T1+FLAIR suffered a significant drop in performance on Philips scanners. The performance decrease on Philips and GE scanners likely reflects a larger uncertainty with smaller number of scans in those categories; inconsistent performance in some models, despite diverse training data, may similarly stem from scanner representation in the training sets (28,31). The nnU-Net T1+FLAIR model had much lower performance on Philips scanners specifically, though it is unclear where this drop in performance originates from given that the model was also trained on images taken from a Philips scanner (28). It is also possible that the images from the Philips scanner category had other features that were difficult for the model to segment.

Furthermore, almost all models performed disproportionately worse on subjects with low PVS severity compared to high-burden cases. This likely reflects training dataset bias, as models used in this study were frequently trained on severe cases to maximize voxel-level representation (23,28,31). For example, mcPVS-Net, trained specifically on high-PVS-burden images, performed significantly better on higher-severity categories (Figure 4, Supplementary Table S8, S9) compared to low-severity images. Consequently, when deployed on heterogeneous cohorts containing some healthy individuals, these models may underperform, potentially skewing longitudinal analyses tracking early appearance of PVS burden.

### 4.3. Clinical Validity

Since these models are ultimately intended for clinical research, they must reliably reproduce biological relationships found via manual delineation. In our ground-truth data, higher PVS counts were significantly associated with Aβ positivity. Strikingly, only the two models specifically trained on AD cohorts (ADNI-PVS-Net and mcPVS-Net) replicated this relationship. Although MedNet-PVS was trained on a clinical cohort and generalized best out-of-distribution based on segmentation performance, it failed to reproduce the significant manual-label relationships. This is arguably the most critical finding of this study: using unvalidated segmentation models in specific disease cohorts risks masking true biological signals, leading to false-negative scientific discoveries (Type II errors).

In the BG, the significant relationships could not be reproduced by any of the models; as also indicated by the regional performance, this reflects greater uncertainty in this region. On the other hand, the negative findings were successfully replicated by other models, with relatively similar effect sizes across the models that were trained with clinical cohorts, namely ADNI-PVS-Net, mcPVS-Net, and MedNet-PVS.

### 4.4. Limitations

This study has several limitations. First, even with rigorous out-of-fold cross-validation, ADNI-PVS-Net retains an inherent domain advantage on the ADNI-3 cohort, so its performance represents an ideal internal scenario rather than true external generalizability. Subjective rater variability in manual ground-truth labeling across the training datasets of the five models also likely accounts for some baseline performance variations. Assessments of robustness between scanner manufacturers may have also been influenced by an unequal number of scanner types in the dataset (Siemens n=39, Philips n=8, GE n=13). In addition, the cross-sectional nature of the data precluded the assessment of longitudinal stability, which is essential for tracking glymphatic dysfunction. Lastly, although the delineation of PVS closely followed the criteria outlined in STRIVE-2 (3,24), was done in conjunction with FLAIR images, and WMH segmentation masks were included, overlap is still possible (3), adding variability in the ground truth set.

### 4.5. Future directions

These results strongly suggest validating off-the-shelf segmentation tools on a subset of their specific cohort before proceeding with population-level analyses. Future studies should evaluate the clinical utility of these models in other high-vascular-burden cohorts in addition to AD, such as vascular dementia and multiple sclerosis, across higher field strengths (7T), and across longitudinal data. Based on the results of this and similar studies (4,19,20,29,32,44), further improvement of PVS segmentation models may benefit from training and benchmarking larger and more diverse datasets, such as through computational challenges or other data sharing methods. For AD specifically, it may be beneficial to train a domain-specialized model for maximum clinical utility. Future nnU-Net-based development should also account for its reliance on automated preprocessing, which may reduce out-of-distribution generalizability. Extrapolating from the segmentation performance, a model built on the newest nnU-Net methods (MedNeXt or nnU-Netv2 with residual encoders (26,44)), trained on T1 and FLAIR, and on AD-specific data with a wide range of PVS burden may achieve higher segmentation performance and better clinical utility for AD.

Taken together, these findings offer a cautionary tale for the field: model choice meaningfully shapes both segmentation accuracy and downstream clinical inferences about PVS burden in AD, underscoring the need for cohort-specific validation in deep learning methods for neuroimaging.

## Data and code availability statement

Data, including clinical, biofluid, genetic, imaging, and neuropathology results from ADNI participants, are shared with approved researchers through the LONI Image and Data Archive (IDA), a secure research data repository. Segmentation data used in this study may be available upon request. Models used in this study are available through the following Github repository: https://github.com/PVS-segmentation-repository

## CRediT author statement

ST: Conceptualization, Data curation, Formal analysis, Investigation, Methodology, Software, Visualization, Validation, Writing – original draft, Writing – review & editing. KY: Supervision, Validation, Writing – review & editing, Methodology. EP: Validation. IH: Writing – review & editing, Formal analysis. PT: Writing – review & editing. JL: Methodology, Supervision, Writing – review & editing. DT: Funding acquisition, Investigation, Project administration, Resources, Supervision, Formal analysis, Writing – review & editing, Data curation, Methodology, Validation.

## Acknowledgments

Data collection and sharing for the Alzheimer’s Disease Neuroimaging Initiative (ADNI) is funded by the National Institute on Aging (National Institutes of Health Grant U19 AG024904). The grantee organization is the Northern California Institute for Research and Education. In the past, ADNI has also received funding from the National Institute of Biomedical Imaging and Bioengineering, the Canadian Institutes of Health Research, and private sector contributions through the Foundation for the National Institutes of Health (FNIH) including generous contributions from the following: AbbVie, Alzheimer’s Association; Alzheimer’s Drug Discovery Foundation; Araclon Biotech; BioClinica, Inc.; Biogen; Bristol-Myers Squibb Company; CereSpir, Inc.; Cogstate; Eisai Inc.; Elan Pharmaceuticals, Inc.; Eli Lilly and Company; EuroImmun; F. Hoffmann-La Roche Ltd and its affiliated company Genentech, Inc.; Fujirebio; GE Healthcare; IXICO Ltd.; Janssen Alzheimer Immunotherapy Research & Development, LLC.; Johnson & Johnson Pharmaceutical Research &Development LLC.; Lumosity; Lundbeck; Merck & Co., Inc.; Meso Scale Diagnostics, LLC.; NeuroRx Research; Neurotrack Technologies; Novartis Pharmaceuticals Corporation; Pfizer Inc.; Piramal Imaging; Servier; Takeda Pharmaceutical Company; and Transition Therapeutics.

## References

1. Iliff JJ, Wang M, Liao Y, Plogg BA, Peng W, Gundersen GA, et al. A Paravascular Pathway Facilitates CSF Flow Through the Brain Parenchyma and the Clearance of Interstitial Solutes, Including Amyloid β. Sci Transl Med. 2012 Aug 15;4(147). doi:10.1126/scitranslmed.3003748

2. Wardlaw JM, Smith EE, Biessels GJ, Cordonnier C, Fazekas F, Frayne R, et al. Neuroimaging standards for research into small vessel disease and its contribution to ageing and neurodegeneration. Lancet Neurol. 2013 Aug;12(8):822–38. doi:10.1016/S1474-4422(13)70124-8

3. Duering M, Biessels GJ, Brodtmann A, Chen C, Cordonnier C, De Leeuw FE, et al. Neuroimaging standards for research into small vessel disease—advances since 2013. Lancet Neurol. 2023 Jul;22(7):602–18. doi:10.1016/S1474-4422(23)00131-X

4. Waymont JMJ, Valdés Hernández M del C, Bernal J, Duarte Coello R, Brown R, Chappell FM, et al. Systematic review and meta-analysis of automated methods for quantifying enlarged perivascular spaces in the brain. NeuroImage. 2024 Aug 15;297:120685. doi:10.1016/j.neuroimage.2024.120685

5. Wardlaw JM, Benveniste H, Nedergaard M, Zlokovic BV, Mestre H, Lee H, et al. Perivascular spaces in the brain: anatomy, physiology and pathology. Nat Rev Neurol. 2020 Mar 15;16(3):137–53. doi:10.1038/s41582-020-0312-z

6. Tang S, Thropp P, Hausle I, Younes K, Tosun D. Spatial coupling of enlarged perivascular spaces and white matter lesions across the Alzheimer’s disease continuum. Front Neurosci. 2026 Apr 1;20:1772024. doi:10.3389/fnins.2026.1772024

7. Menze I, Bernal J, Kaya P, Aki Ç, Pfister M, Geisendörfer J, et al. Perivascular space enlargement accelerates in ageing and Alzheimer’s disease pathology: evidence from a three-year longitudinal multicentre study. Alzheimers Res Ther. 2024 Oct 31;16(1):242. doi:10.1186/s13195-024-01603-8

8. Ong JJH, Leow YJ, Qiu B, Tanoto P, Zailan FZ, Sandhu GK, et al. Association of Enlarged Perivascular Spaces With Early Serum and Neuroimaging Biomarkers of Alzheimer Disease Pathology. Neurology. 2025 Sep 23;105(6):e213836. doi:10.1212/WNL.0000000000213836

9. Ishida K, Yamada K, Nishiyama R, Hashimoto T, Nishida I, Abe Y, et al. Glymphatic system clears extracellular tau and protects from tau aggregation and neurodegeneration. J Exp Med. 2022 Mar 7;219(3):e20211275. doi:10.1084/jem.20211275

10. Dagum P, Elbert DL, Giovangrandi L, Singh T, Venkatesh VV, Corbellini A, et al. The glymphatic system clears amyloid beta and tau from brain to plasma in humans [Internet]. Neurology; 2024 [cited 2026 Sep 10]. Available from: http://medrxiv.org/lookup/doi/10.1101/2024.07.30.24311248 doi:10.1101/2024.07.30.24311248

11. Yamamoto EA, Bagley JH, Geltzeiler M, Sanusi OR, Dogan A, Liu JJ, et al. The perivascular space is a conduit for cerebrospinal fluid flow in humans: A proof-of-principle report. Proc Natl Acad Sci. 2024 Oct 15;121(42):e2407246121. doi:10.1073/pnas.2407246121

12. Wang ML, Zou QQ, Sun Z, Wei XE, Li PY, Wu X, et al. Associations of MRI-visible perivascular spaces with longitudinal cognitive decline across the Alzheimer’s disease spectrum. Alzheimers Res Ther. 2022 Dec 13;14(1):185. doi:10.1186/s13195-022-01136-y

13. Hong H, Hong L, Luo X, Zeng Q, Li K, Wang S, et al. The relationship between amyloid pathology, cerebral small vessel disease, glymphatic dysfunction, and cognition: a study based on Alzheimer’s disease continuum participants. Alzheimers Res Ther. 2024 Feb 20;16(1):43. doi:10.1186/s13195-024-01407-w

14. Kim HJ, Cho H, Park M, Kim JW, Ahn SJ, Lyoo CH, et al. MRI-Visible Perivascular Spaces in the Centrum Semiovale Are Associated with Brain Amyloid Deposition in Patients with Alzheimer Disease–Related Cognitive Impairment. Am J Neuroradiol. 2021 Jul;42(7):1231–8. doi:10.3174/ajnr.A7155

15. Sepehrband F, Barisano G, Sheikh-Bahaei N, Choupan J, Cabeen RP, Lynch KM, et al. Volumetric distribution of perivascular space in relation to mild cognitive impairment. Neurobiol Aging. 2021 Mar;99:28–43. doi:10.1016/j.neurobiolaging.2020.12.010

16. Banerjee G, Kim HJ, Fox Z, Jäger HR, Wilson D, Charidimou A, et al. MRI-visible perivascular space location is associated with Alzheimer’s disease independently of amyloid burden. Brain. 2017 Apr 1;140(4):1107–16. doi:10.1093/brain/awx003

17. Jeong SH, Cha J, Park M, Jung JH, Ye BS, Sohn YH, et al. Association of Enlarged Perivascular Spaces With Amyloid Burden and Cognitive Decline in Alzheimer Disease Continuum. Neurology. 2022 Oct 18;99(16). doi:10.1212/WNL.0000000000200989

18. Younes K, Cobbigo Y, Tsuie T, Wang E, Wolf A, Joie RL, et al. Divergent enlarged perivascular spaces volumes in early versus late age-of-onset Alzheimer’s disease [Internet]. 2023 [cited 2024 May 8]. Available from: http://medrxiv.org/lookup/doi/10.1101/2023.08.01.23293514 doi:10.1101/2023.08.01.23293514

19. Dash S, del C. Valdés Hernández M, Pham W, Salah Khlif M, Brodtmann A, Joliot M, et al. Assessing Generalisation of Perivascular Space Segmentation Across Heterogeneous MRI Cohorts: The DoRA-PVS Challenge 2026. In: Ni H, Cafolla D, editors. Artificial Intelligence in Healthcare. Cham: Springer Nature Switzerland; 2027. p. 391–404. doi:10.1007/978-3-032-35393-1_28

20. Wu Y, Zhang Y, Dong Z, Ji F, Tan AS, Tan G, et al. Standardized evaluation of automatic methods for perivascular spaces segmentation in MRI – MICCAI 2024 challenge results. Med Image Anal. 2026 Nov;114:104227. doi:10.1016/j.media.2026.104227

21. Pham W, Lynch M, Spitz G, O’Brien T, Vivash L, Sinclair B, et al. A critical guide to the automated quantification of perivascular spaces in magnetic resonance imaging. Front Neurosci. 2022 Dec 14;16:1021311. doi:10.3389/fnins.2022.1021311

22. Moses J, Sinclair B, Law M, O’Brien TJ, Vivash L. Automated Methods for Detecting and Quantitation of Enlarged Perivascular spaces on MRI. J Magn Reson Imaging. 2023 Jan;57(1):11–24. doi:10.1002/jmri.28369

23. Huang P, Liu L, Zhang Y, Zhong S, Liu P, Hong H, et al. Development and validation of a perivascular space segmentation method in multi-center datasets. NeuroImage. 2024 Sep;298:120803. doi:10.1016/j.neuroimage.2024.120803

24. Potter GM, Chappell FM, Morris Z, Wardlaw JM. Cerebral Perivascular Spaces Visible on Magnetic Resonance Imaging: Development of a Qualitative Rating Scale and its Observer Reliability. Cerebrovasc Dis. 2015;39(3–4):224–31. doi:10.1159/000375153

25. Adams HHH, Cavalieri M, Verhaaren BFJ, Bos D, Van Der Lugt A, Enzinger C, et al. Rating Method for Dilated Virchow–Robin Spaces on Magnetic Resonance Imaging. Stroke. 2013 Jun;44(6):1732–5. doi:10.1161/STROKEAHA.111.000620

26. Isensee F, Jaeger PF, Kohl SAA, Petersen J, Maier-Hein KH. nnU-Net: a self-configuring method for deep learning-based biomedical image segmentation. Nat Methods. 2021 Feb;18(2):203–11. doi:10.1038/s41592-020-01008-z

27. Isensee F, Wald T, Ulrich C, Baumgartner M, Roy S, Maier-Hein K, et al. nnU-Net Revisited: A Call for Rigorous Validation in 3D Medical Image Segmentation [Internet]. arXiv; 2024 [cited 2026 Sep 10]. Available from: http://arxiv.org/abs/2404.09556 10.48550/arXiv.2404.09556

28. Pham W, Jarema A, Rim D, Chen Z, Khlif M, Macefield V, et al. A comprehensive framework for automated segmentation of perivascular spaces in brain MRI with the nnU-Net. Neuroradiology. 2026 Jun;68(6):1465–83. doi:10.1007/s00234-026-03993-y

29. Sinclair B, Pham W, Vivash L, Moses J, Lynch M, Dorfman K, et al. Perivascular space identification nnUNet for generalised usage (PINGU). Med Image Anal. 2026 Mar;109:103903. doi:10.1016/j.media.2025.103903

30. LeFevre JD, Robb WH, Liu D, Jackson TB, Pechman KR, Shashikumar N, et al. A comparative evaluation of multiple enlarged perivascular space segmentation tools. Magn Reson Imaging. 2026 Dec;134:110749. doi:10.1016/j.mri.2026.110749

31. Low ZXB, Zhang R, Min H, Pham W, Vivash L, Moses J, et al. MedNet-PVS: A MedNeXt-Based Deep Learning Model for Automated Segmentation of Perivascular Spaces. arXiv. 2025. 10.48550/arXiv.2508.20256

32. Bitar L, Díaz M, Coello RD, Valdés-Hernández M d.C., Mattern H, Neumann K, et al. DRIPS: Domain Randomisation for Image-based Perivascular spaces Segmentation. medRxiv. 2025 Oct 30;2025.10.22.25337423. doi:10.1101/2025.10.22.25337423 PubMed PMID: 41282908; PubMed Central PMCID: PMC12633610.

33. Gibson E, Ramirez J, Woods LA, Berberian S, Ottoy J, Scott CJM, et al. segcsvdPVS : A Convolutional Neural Network-Based Tool for Quantification of Enlarged Perivascular Spaces (PVS) on T1 -Weighted Images. Hum Brain Mapp. 2026 Feb 1;47(2):e70462. doi:10.1002/hbm.70462

34. Kang J, Bak D, Shin N, Kim HG, Nam Y. Improved BG - PVS Quantification in Infant Brain MRI Using Anatomy-Informed Pseudo-Labels for Joint BG and PVS Segmentation. J Magn Reson Imaging. 2026 Jul;64(1):168–83. doi:10.1002/jmri.70298

35. Sudre CH, Van Wijnen K, Dubost F, Adams H, Atkinson D, Barkhof F, et al. Where is VALDO? VAscular Lesions Detection and segmentatiOn challenge at MICCAI 2021. Med Image Anal. 2024 Jan;91:103029. doi:10.1016/j.media.2023.103029

36. Liu S, Hou B, You H, Zhang Y, Zhu Y, Ma C, et al. The Association Between Perivascular Spaces and Cerebral Blood Flow, Brain Volume, and Cardiovascular Risk. Front Aging Neurosci. 2021 Aug 31;13:599724. doi:10.3389/fnagi.2021.599724

37. Liu W, Bu W, Meng L, Luo W, Liu C, Li X, et al. Spatial distribution of enlarged perivascular spaces as a potential biomarker for distinguishing vascular dementia from Alzheimer’s disease in older adults. Clin Neurol Neurosurg. 2025 Oct;257:109098. doi:10.1016/j.clineuro.2025.109098

38. Jack Jr. CR, Arani A, Borowski BJ, Cash DM, Crawford K, Das SR, et al. Overview of ADNI MRI. Alzheimers Dement. 2024;20(10):7350–60. doi:10.1002/alz.14166

39. Tillin T, Hughes AD, Mayet J, Whincup P, Sattar N, Forouhi NG, et al. The Relationship Between Metabolic Risk Factors and Incident Cardiovascular Disease in Europeans, South Asians, and African Caribbeans. J Am Coll Cardiol. 2013 Apr;61(17):1777–86. doi:10.1016/j.jacc.2012.12.046

40. Ikram MA, Brusselle G, Ghanbari M, Goedegebure A, Ikram MK, Kavousi M, et al. Objectives, design and main findings until 2020 from the Rotterdam Study. Eur J Epidemiol. 2020 May;35(5):483–517. doi:10.1007/s10654-020-00640-5

41. Koo TK, Li MY. A Guideline of Selecting and Reporting Intraclass Correlation Coefficients for Reliability Research. J Chiropr Med. 2016 Jun;15(2):155–63. doi:10.1016/j.jcm.2016.02.012

42. Schmidt P. (2017). Bayesian inference for structured additive regression models for large-scale problems with applications to medical imaging (PhD thesis). Ludwig Maximilians-Universität München. Available online at: http://nbn-resolving.de/urn:nbn:de:bvb:19-203731

43. Schmidt P, Gaser C, Arsic M, Buck D, Förschler A, Berthele A, et al. An automated tool for detection of FLAIR-hyperintense white-matter lesions in Multiple Sclerosis. NeuroImage. 2012 Feb;59(4):3774–83. doi:10.1016/j.neuroimage.2011.11.032

44. Roy S, Koehler G, Ulrich C, Baumgartner M, Petersen J, Isensee F, et al. MedNeXt: Transformer-driven Scaling of ConvNets for Medical Image Segmentation [Internet]. arXiv; 2024 [cited 2026 Sep 10]. Available from: http://arxiv.org/abs/2303.09975 10.48550/arXiv.2303.09975

45. Maier-Hein L, Reinke A, Godau P, Tizabi MD, Buettner F, Christodoulou E, et al. Metrics reloaded: recommendations for image analysis validation. Nat Methods. 2024 Feb;21(2):195–212. doi:10.1038/s41592-023-02151-z

46. Gibson E, Ramirez J, Woods LA, Berberian S, Ottoy J, Scott CJM, et al. segcsvdPVS : A Convolutional Neural Network-Based Tool for Quantification of Enlarged Perivascular Spaces (PVS) on T1 -Weighted Images. Hum Brain Mapp. 2026 Feb 5;47(2):e70462. doi:10.1002/hbm.70462 PubMed PMID: 41641899; PubMed Central PMCID: PMC12874998.

47. Seabold S, Perktold J. Statsmodels: Econometric and Statistical Modeling with Python. In. Austin, Texas; 2010. p. 92–6. doi:10.25080/Majora-92bf1922-011

