## Supplementary material for "Examining the Clinical Utility of Deep Learning Models for Perivascular Space Segmentation in Alzheimer’s Disease": supp file

**Supplementary Materials**

Figures

*Figure S1: (A) Illustration of white matter hyperintensity in example FLAIR image (top) and corresponding segmentation (bottom); (B) example slab segments that were used for SABRE in the VALDO dataset, and (C) example slice and regional segments that were used for RSS.*

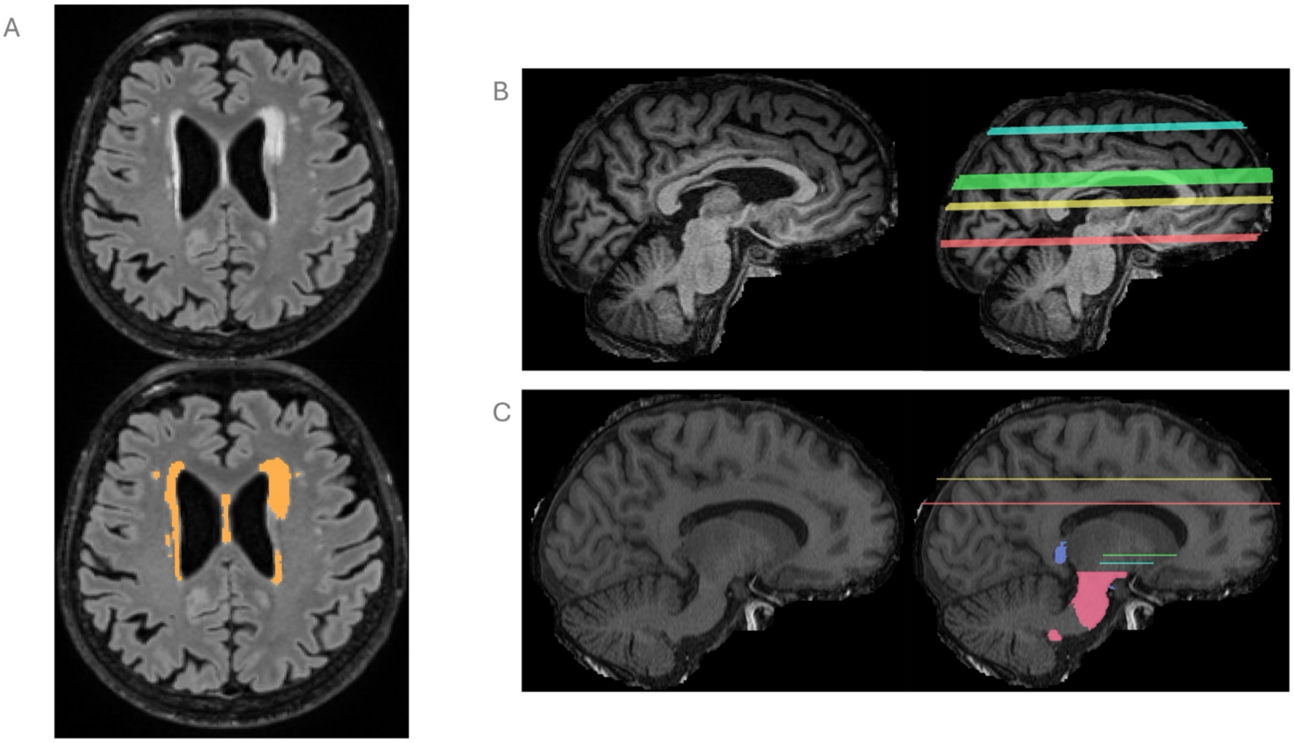

*Figure S2: Table with figures illustrating how overall, object-level, voxel-wise, and lesion-wise Dice are calculated, including score for example figure and method of aggregation over multiple subjects.*

| Metric Name | Image Example | Total TP, FP, FN | Score for the example | Method of aggregation over all samples |
|---|---|---|---|---|
| Overall Dice | 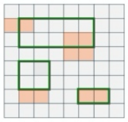 | TP = 5<br>FP = 5<br>FN = 11 | = 0.23 | Avg (∑(TP, FP, FN) over image) over subjects |
| Object-level Dice | 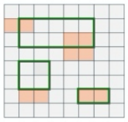 | TP = 2<br>FP = 1<br>FN = 1 | = 0.5 | Avg (∑(TP, FP, FN) over image) over subjects |
| Voxel-wise Dice | 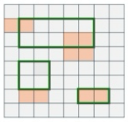 | TP = 5<br>FP = 3<br>FN = 7 | = 0.33 | Avg (∑(TP, FP, FN) over image) over subjects |
| Lesion-wise Dice | 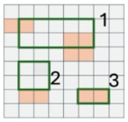 | 1: TP = 3, FN = 7, FP = 3<br>2: N/A<br>3: TP = 2, FN = 0, FP = 0 | = (0.23 + 1) / 2<br>= 0.62 | Avg (avg lesion over image) over subjects |

Tables

Table 1: List of nnU-Net based PVS segmentation models

| Research Study | Model or Team name | Segmentation of WM/BG? | Requires inputs other than T1 or T1+FLAIR? | Publicly Available? | Include? |
|---|---|---|---|---|---|
| Sinclair et al 2025 (1) | PINGU | Yes | No | Yes | No* |
| Huang et al 2025 (2) | mcPVS-Net | Yes | No | Yes | Yes |
| Pham et al 2026 (3) | nnU-Net T1 / nnU-Net T1+FLAIR | Yes | No | Yes | Yes |
| Low et al 2025 (4) | MedNet-PVS | Yes | No | Yes | Yes |
| LeFevre et al 2026 (5) | DORES | Yes | No | No | Yes |
| Hürter et al 2026. (6) | - | Yes | Yes | Yes | No |
| Wu et al 2026 — MICCAI 2024 Challenge (7) | bear_walker | Yes | Yes | Yes | No |
| | nic-vicorob | Yes | Yes | Yes | No |
| | BIG_AEHRC | Yes | Yes | Yes | No |
| Dash et al 2026 — AliH DoRA-PVS Challenge (8) | WAVM | Yes | No | No | No |
| | VICOROBIGR | Yes | No | No | No |

*Table 1: Review of published nnU-Net models for PVS segmentation. Models were only included if they were publicly available (at the time of this study), did not require more than either T1 or T1+FLAIR images as input, and included segmentation of both WM / CSO regions and BG regions.*
**Although PINGU fits the criteria for inclusion, it shares training data with MedNet-PVS but uses an older version of nnU-Net (nnunetv1) and was therefore excluded for redundancy.*

*Model references:*


1. *Sinclair B, Pham W, Vivash L, Moses J, Lynch M, Dorfman K, et al. Perivascular space identification nnUNet for generalised usage (PINGU). Med Image Anal. 2026 Mar;109:103903. doi:10.1016/j.media.2025.103903*
2. *Huang P, Liu L, Zhang Y, Zhong S, Liu P, Hong H, et al. Development and validation of a perivascular space segmentation method in multi-center datasets. NeuroImage. 2024 Sep;298:120803. doi:10.1016/j.neuroimage.2024.120803*
3. *Pham W, Jarema A, Rim D, Chen Z, Khlif M, Macefield V, et al. A comprehensive framework for automated segmentation of perivascular spaces in brain MRI with the nnU-Net. Neuroradiology. 2026 Jun;68(6):1465–83. doi:10.1007/s00234-026-03993-y*
4. *Low ZXB, Zhang R, Min H, Pham W, Vivash L, Moses J, et al. MedNet-PVS: A MedNeXt-Based Deep Learning Model for Automated Segmentation of Perivascular Spaces. arXiv. 2025. doi:https://doi.org/10.48550/arXiv.2508.20256*
5. *LeFevre JD, Robb WH, Liu D, Jackson TB, Pechman KR, Shashikumar N, et al. A comparative evaluation of multiple enlarged perivascular space segmentation tools. Magn Reson Imaging. 2026 Dec;134:110749. doi:10.1016/j.mri.2026.110749*
6. *Hürter NM, Schmenger VS, Barda T, Thalhammer M. Higher perivascular space volume in very premature born adults. Available from: medRxiv 2026.05.23.26353943; doi: https://doi.org/10.64898/2026.05.23.26353943*
7. *Wu Y, Zhang Y, Dong Z, Ji F, Tan AS, Tan G, et al. Standardized evaluation of automatic methods for perivascular spaces segmentation in MRI – MICCAI 2024 challenge results. Med Image Anal. 2026 Nov;114:104227. doi:10.1016/j.media.2026.104227*
8. *Dash S, del C. Valdés Hernández M, Pham W, Salah Khlif M, Brodtmann A, Joliot M, et al. Assessing Generalisation of Perivascular Space Segmentation Across Heterogeneous MRI Cohorts: The DoRA-PVS Challenge 2026. In: Ni H, Cafolla D, editors. Artificial Intelligence in Healthcare. Cham: Springer Nature Switzerland; 2027. p. 391–404. doi:10.1007/978-3-032-35393-1_28*


Table 2: Overall performance on ADNI-3 (n=60)

| Model | Overall | Object-level | Voxel-wise | Lesion-wise | Count r | Count CCC | Vol r | Vol CCC |
|---|---|---|---|---|---|---|---|---|

| | | | | | | | | |
|---|---|---|---|---|---|---|---|---|
| mcPVS-Net | $0.24 \pm 0.13^{bde}$ | $0.30 \pm 0.15^{be}$ | $0.50 \pm 0.10^{bde}$ | $0.76 \pm 0.19^{bce}$ | r=0.83; p<0.001 | 0.28 | r=0.79; p<0.001 | 0.3 |
| nnU-Net T1 | $0.27 \pm 0.14^{ae}$ | $0.34 \pm 0.16^{acde}$ | $0.56 \pm 0.10^{ae}$ | $0.54 \pm 0.23^{acd}$ | r=0.79; p<0.001 | 0.5 | r=0.76; p<0.001 | 0.49 |
| nnU-Net T1+FLAIR | $0.23 \pm 0.16^{e}$ | $0.26 \pm 0.19^{be}$ | $0.49 \pm 0.21^{e}$ | $0.33 \pm 0.27^{abde}$ | r=0.54; p<0.001 | 0.47 | r=0.55; p<0.001 | 0.48 |
| MedNet-PVS | $0.29 \pm 0.15^{ae}$ | $0.29 \pm 0.17^{be}$ | $0.57 \pm 0.11^{ae}$ | $0.79 \pm 0.16^{bce}$ | r=0.75; p<0.001 | 0.2 | r=0.70; p<0.001 | 0.32 |
| ADNI-PVS-Net | $0.58 \pm 0.14^{abcd}$ | $0.66 \pm 0.14^{abcd}$ | $0.73 \pm 0.12^{abcd}$ | $0.53 \pm 0.18^{acd}$ | r=0.92; p<0.001 | 0.86 | r=0.92; p<0.001 | 0.61 |

Table 3: Overall performance on VALDO (split by dataset, n=6 per dataset)

| Model | Overall | Object-level | Voxel-wise | Lesion-wise | Count r | Count CCC | Vol r | Vol CCC |
|---|---|---|---|---|---|---|---|---|
| RSS (1.5T) | | | | | | | | |
| mcPVS-Net | $0.27 \pm 0.14^{ce}$ | $0.45 \pm 0.12^{ce}$ | $0.52 \pm 0.16^{c}$ | $0.27 \pm 0.14^{cde}$ | r=0.77; p=0.07 | 0.78 | r=0.83; p=0.04 | 0.53 |
| nnU-Net T1 | $0.15 \pm 0.09^{cd}$ | $0.31 \pm 0.09^{d}$ | $0.49 \pm 0.11$ | $0.18 \pm 0.10^{cd}$ | r=-0.71; p=0.11 | -0.45 | r=-0.77; p=0.07 | -0.07 |
| nnU-Net T1+FLAIR | $0.06 \pm 0.05^{abd}$ | $0.21 \pm 0.14^{ad}$ | $0.39 \pm 0.21^{a}$ | $0.07 \pm 0.05^{abd}$ | r=0.66; p=0.16 | 0.07 | r=0.71; p=0.11 | 0.01 |
| MedNet-PVS | $0.30 \pm 0.09^{bce}$ | $0.49 \pm 0.10^{bc}$ | $0.55 \pm 0.13$ | $0.38 \pm 0.12^{abce}$ | r=0.66; p=0.16 | 0.61 | r=0.83; p=0.04 | 0.40 |
| ADNI-PVS-Net | $0.11 \pm 0.07^{ad}$ | $0.25 \pm 0.14^{a}$ | $0.42 \pm 0.21$ | $0.09 \pm 0.05^{ad}$ | r=0.77; p=0.07 | 0.12 | r=0.77; p=0.07 | 0.06 |
| SABRE (3T) | | | | | | | | |
| mcPVS-Net | $0.22 \pm 0.10^{e}$ | $0.26 \pm 0.11^{ce}$ | $0.46 \pm 0.07$ | $0.14 \pm 0.06^{ce}$ | r=0.09; p=0.87 | 0.01 | r=0.49; p=0.33 | 0.09 |
| nnU-Net T1 | $0.17 \pm 0.06^{cde}$ | $0.17 \pm 0.08^{cde}$ | $0.47 \pm 0.11$ | $0.09 \pm 0.07^{cde}$ | r=-0.09; p=0.87 | 0.01 | r=0.09; p=0.87 | 0.04 |
| nnU-Net T1+FLAIR | $0.07 \pm 0.05^{bd}$ | $0.08 \pm 0.06^{abd}$ | $0.46 \pm 0.14$ | $0.05 \pm 0.03^{abd}$ | r=-0.66; p=0.16 | -0.02 | r=-0.09; p=0.87 | -0.01 |
| MedNet-PVS | $0.26 \pm 0.06^{bce}$ | $0.29 \pm 0.09^{bce}$ | $0.51 \pm 0.08$ | $0.17 \pm 0.07^{bce}$ | r=0.41; p=0.42 | 0.12 | r=0.66; p=0.16 | 0.13 |
| ADNI-PVS-Net | $0.04 \pm 0.03^{abd}$ | $0.06 \pm 0.05^{abd}$ | $0.35 \pm 0.20$ | $0.02 \pm 0.01^{abd}$ | r=0.49; p=0.33 | 0.01 | r=0.60; p=0.21 | 0.01 |

Table 4: Performance by brain region

| Model | Overall | Object-level | Voxel-wise | Lesion-wise | Count r | Count CCC | Vol r | Vol CCC |
|---|---|---|---|---|---|---|---|---|
| White matter | | | | | | | | |
| mcPVS-Net | $0.21 \pm 0.12^{bde}$ | $0.26 \pm 0.15^{be}$ | $0.49 \pm 0.13^{bde}$ | $0.69 \pm 0.20^{bce}$ | r=0.84; p<0.001 | 0.28 | r=0.84; p<0.001 | 0.31 |
| nnU-Net T1 | $0.25 \pm 0.14^{ae}$ | $0.30 \pm 0.17^{ade}$ | $0.55 \pm 0.14^{ae}$ | $0.48 \pm 0.24^{acd}$ | r=0.77; p<0.001 | 0.52 | r=0.78; p<0.001 | 0.54 |
| nnU-Net T1+FLAIR | $0.23 \pm 0.17^{e}$ | $0.26 \pm 0.19^{e}$ | $0.47 \pm 0.24^{de}$ | $0.33 \pm 0.26^{abde}$ | r=0.59; p<0.001 | 0.54 | r=0.61; p<0.001 | 0.54 |
| MedNet-PVS | $0.26 \pm 0.16^{ae}$ | $0.26 \pm 0.16^{be}$ | $0.57 \pm 0.15^{ace}$ | $0.69 \pm 0.21^{bce}$ | r=0.75; p<0.001 | 0.2 | r=0.73; p<0.001 | 0.35 |
| ADNI-PVS-Net | $0.59 \pm 0.17^{abcd}$ | $0.65 \pm 0.18^{abcd}$ | $0.73 \pm 0.16^{abcd}$ | $0.55 \pm 0.21^{acd}$ | r=0.94; p<0.001 | 0.88 | r=0.94; p<0.001 | 0.66 |
| Basal Ganglia | | | | | | | | |
| mcPVS-Net | $0.20 \pm 0.15^{ce}$ | $0.27 \pm 0.18^{bce}$ | $0.47 \pm 0.25^{ce}$ | $0.36 \pm 0.26^{bcd}$ | r=0.67; p<0.001 | 0.53 | r=0.53; p<0.001 | 0.61 |
| nnU-Net T1 | $0.20 \pm 0.17^{ce}$ | $0.22 \pm 0.15^{ace}$ | $0.45 \pm 0.25^{ce}$ | $0.26 \pm 0.21^{acde}$ | r=0.46; p<0.001 | 0.31 | r=0.45; p<0.001 | 0.37 |
| nnU-Net T1+FLAIR | $0.11 \pm 0.15^{abde}$ | $0.12 \pm 0.15^{abde}$ | $0.26 \pm 0.27^{abde}$ | $0.11 \pm 0.16^{abde}$ | r=-0.01; p=0.96 | -0.0 | r=-0.01; p=0.93 | -0.02 |
| MedNet-PVS | $0.22 \pm 0.16^{ce}$ | $0.25 \pm 0.17^{ce}$ | $0.47 \pm 0.24^{ce}$ | $0.44 \pm 0.29^{abc}$ | r=0.50; p<0.001 | 0.43 | r=0.57; p<0.001 | 0.57 |
| ADNI-PVS-Net | $0.40 \pm 0.27^{abcd}$ | $0.50 \pm 0.30^{abcd}$ | $0.56 \pm 0.31^{abcd}$ | $0.37 \pm 0.28^{bc}$ | r=0.80; p<0.001 | 0.7 | r=0.85; p<0.001 | 0.53 |

Table 5: Performance by WMH region

| Model | Overall | Object-level | Voxel-wise | Lesion-wise | False Discovery Rate (WMH) | FP Rate (per WMH Vol.) |
|---|---|---|---|---|---|---|
| mcPVS-Net | $0.20 \pm 0.21^{e}$ | $0.26 \pm 0.23$ | $0.58 \pm 0.34^{c}$ | $0.62 \pm 0.42^{bce}$ | $0.79 \pm 0.23^{ce}$ | $21.79 \pm 16.91^{cde}$ |
| nnU-Net T1 | $0.16 \pm 0.25^{e}$ | $0.24 \pm 0.27$ | $0.47 \pm 0.38$ | $0.51 \pm 0.43^{acde}$ | $0.75 \pm 0.31^{ce}$ | $19.43 \pm 18.54^{cde}$ |
| nnU-Net T1+FLAIR | $0.16 \pm 0.27^{e}$ | $0.22 \pm 0.29^{e}$ | $0.37 \pm 0.40^{ad}$ | $0.42 \pm 0.46^{abd}$ | $0.43 \pm 0.42^{abde}$ | $11.73 \pm 17.05^{abde}$ |
| MedNet-PVS | $0.18 \pm 0.22^{e}$ | $0.26 \pm 0.23$ | $0.62 \pm 0.33^{c}$ | $0.72 \pm 0.39^{bce}$ | $0.81 \pm 0.20^{ce}$ | $32.53 \pm 26.41^{abce}$ |

| | | | | | | |
|---|---|---|---|---|---|---|
| ADNI-PVS-Net | 0.38 ± 0.38[abcd] | 0.43 ± 0.40[c] | 0.56 ± 0.45 | 0.36 ± 0.38[abd] | 0.13 ± 0.30[abcd] | 1.04 ± 2.80[abcd] |

*Table 2-5: Values are mean ± SD. Superscript letters denote a statistically significant difference (paired Wilcoxon signed-rank test, FDR-corrected across the 10 pairwise comparisons within each region/metric family, $p < 0.05$) from the model indicated by that letter.*
*Note for Table S3: Due to the small sample size in this external dataset (n=6 per cohort), FDR correction removed almost all significance. Superscript letters in this table therefore reflect uncorrected (raw) p-values.*
*Key: a = mcPVS-Net; b = nnU-Net T1; c = nnU-Net T1+FLAIR; d = MedNet-PVS; e = ADNI-PVS-Net*

Table 6: Performance by diagnostic category

| Model | Overall | Object-level | Voxel-wise | Lesion-wise |
|---|---|---|---|---|
| CU (n=20) | | | | |
| mcPVS-Net | 0.23 ± 0.12 | 0.28 ± 0.14 | 0.50 ± 0.14 | 0.78 ± 0.21 |
| nnU-Net T1 | 0.25 ± 0.14 | 0.31 ± 0.18 | 0.56 ± 0.15 | 0.56 ± 0.26 |
| nnU-Net T1+FLAIR | 0.24 ± 0.17 | 0.28 ± 0.19 | 0.47 ± 0.25 | 0.40 ± 0.27 |
| MedNet-PVS | 0.24 ± 0.14 | 0.24 ± 0.15 | 0.53 ± 0.15 | 0.81 ± 0.20 |
| ADNI-PVS-Net | 0.56 ± 0.19 | 0.64 ± 0.19 | 0.70 ± 0.18 | 0.50 ± 0.22 |
| MCI (n=20) | | | | |
| mcPVS-Net | 0.24 ± 0.16 | 0.31 ± 0.18 | 0.50 ± 0.07 | 0.75 ± 0.18 |
| nnU-Net T1 | 0.29 ± 0.14 | 0.37 ± 0.17 | 0.54 ± 0.08 | 0.48 ± 0.18 |
| nnU-Net T1+FLAIR | 0.20 ± 0.16 | 0.24 ± 0.19 | 0.47 ± 0.19 | 0.27 ± 0.23 |
| MedNet-PVS | 0.29 ± 0.17 | 0.30 ± 0.20 | 0.57 ± 0.11 | 0.79 ± 0.13 |
| ADNI-PVS-Net | 0.59 ± 0.08 | 0.67 ± 0.10 | 0.75 ± 0.06 | 0.52 ± 0.16 |
| AD (n=20) | | | | |
| mcPVS-Net | 0.24 ± 0.10 | 0.30 ± 0.13 | 0.49 ± 0.07 | 0.75 ± 0.19 |
| nnU-Net T1 | 0.27 ± 0.12 | 0.35 ± 0.14 | 0.57 ± 0.06 | 0.56 ± 0.25 |
| nnU-Net T1+FLAIR | 0.24 ± 0.16 | 0.27 ± 0.19 | 0.52 ± 0.20 | 0.34 ± 0.31 |
| MedNet-PVS | 0.33 ± 0.14 | 0.33 ± 0.15 | 0.60 ± 0.06 | 0.76 ± 0.16 |
| ADNI-PVS-Net | 0.60 ± 0.12 | 0.66 ± 0.11 | 0.75 ± 0.06 | 0.58 ± 0.17 |

*Table 6: Values are mean ± SD. Superscript numbers denote a statistically significant pairwise difference between diagnostic groups (Mann-Whitney U test, FDR-corrected across the 3 pairwise comparisons within each model/metric, $q < 0.05$).*
*Key: 1 = CU vs MCI; 2 = CU vs AD; 3 = MCI vs AD*

Table 7: Performance by scanner type

| Model | Overall | Object-level | Voxel-wise | Lesion-wise |
|---|---|---|---|---|
| SIEMENS (n=39) | | | | |
| mcPVS-Net | 0.23 ± 0.12 | 0.28 ± 0.15 | 0.49 ± 0.11 | 0.74 ± 0.21[1] |
| nnU-Net T1 | 0.27 ± 0.13[1,2] | 0.33 ± 0.15[1,2] | 0.56 ± 0.11 | 0.61 ± 0.21[1] |
| nnU-Net T1+FLAIR | 0.30 ± 0.13[1,2] | 0.34 ± 0.16[1,2] | 0.56 ± 0.13[1] | 0.47 ± 0.23[1,2] |
| MedNet-PVS | 0.27 ± 0.15[2] | 0.26 ± 0.15[2] | 0.56 ± 0.12 | 0.79 ± 0.18[1,2] |
| ADNI-PVS-Net | 0.56 ± 0.15 | 0.63 ± 0.15 | 0.72 ± 0.14 | 0.50 ± 0.18[1] |
| PHILIPS (n=8) | | | | |
| mcPVS-Net | 0.18 ± 0.10 | 0.25 ± 0.09 | 0.48 ± 0.08 | 0.89 ± 0.08[1,3] |
| nnU-Net T1 | 0.13 ± 0.07[1,3] | 0.19 ± 0.12[1,3] | 0.51 ± 0.08 | 0.19 ± 0.13[1,3] |
| nnU-Net T1+FLAIR | 0.03 ± 0.05[1,3] | 0.03 ± 0.06[1,3] | 0.16 ± 0.23[1,3] | 0.03 ± 0.06[1,3] |
| MedNet-PVS | 0.20 ± 0.09[3] | 0.19 ± 0.08[3] | 0.53 ± 0.11 | 0.90 ± 0.06[1,3] |
| ADNI-PVS-Net | 0.65 ± 0.08 | 0.73 ± 0.11 | 0.76 ± 0.05 | 0.72 ± 0.10[1,3] |
| GE (n=13) | | | | |
| mcPVS-Net | 0.30 ± 0.14 | 0.39 ± 0.16 | 0.51 ± 0.07 | 0.74 ± 0.14[3] |
| nnU-Net T1 | 0.36 ± 0.12[2,3] | 0.46 ± 0.13[2,3] | 0.59 ± 0.06 | 0.52 ± 0.16[3] |
| nnU-Net T1+FLAIR | 0.14 ± 0.13[2,3] | 0.19 ± 0.16[2,3] | 0.48 ± 0.23[3] | 0.11 ± 0.11[2,3] |
| MedNet-PVS | 0.39 ± 0.15[2,3] | 0.43 ± 0.17[2,3] | 0.60 ± 0.08 | 0.72 ± 0.12[2,3] |
| ADNI-PVS-Net | 0.60 ± 0.09 | 0.68 ± 0.10 | 0.74 ± 0.06 | 0.53 ± 0.18[3] |

*Table 7: Values are mean ± SD. Superscript numbers denote a statistically significant pairwise difference between scanner types (Mann-Whitney U test, FDR-corrected across the 3 pairwise comparisons within each model/metric, $p < 0.05$).*
*Key: 1 = SIEMENS vs PHILIPS; 2 = SIEMENS vs GE; 3 = PHILIPS vs GE*

Table 8A: Performance by PVS severity categories — White matter

| Model | Overall | Object-level | Voxel-wise | Lesion-wise |
|---|---|---|---|---|
| Rating 1 (n=12) | | | | |
| mcPVS-Net | 0.09 ± 0.07[1,2,3] | 0.09 ± 0.07[1,2,3] | 0.48 ± 0.22 | 0.62 ± 0.27 |

| | | | | |
|---|---|---|---|---|
| nnU-Net T1 | 0.13 ± 0.10[2,3] | 0.14 ± 0.09[1,2,3] | 0.47 ± 0.20 | 0.40 ± 0.30 |
| nnU-Net T1+FLAIR | 0.13 ± 0.15 | 0.15 ± 0.17[2,3] | 0.36 ± 0.29 | 0.26 ± 0.29 |
| MedNet-PVS | 0.12 ± 0.10[1,2,3] | 0.10 ± 0.09[1,2,3] | 0.52 ± 0.24 | 0.63 ± 0.26 |
| ADNI-PVS-Net | 0.54 ± 0.29 | 0.48 ± 0.27[1] | 0.66 ± 0.33 | 0.56 ± 0.33 |
| Rating 2 (n=19) | | | | |
| mcPVS-Net | 0.16 ± 0.09[1,4,5] | 0.19 ± 0.09[1,4,5] | 0.46 ± 0.13 | 0.69 ± 0.23 |
| nnU-Net T1 | 0.21 ± 0.12[4,5] | 0.24 ± 0.12[1,4,5] | 0.54 ± 0.16 | 0.45 ± 0.28 |
| nnU-Net T1+FLAIR | 0.19 ± 0.16 | 0.19 ± 0.16[4,5] | 0.42 ± 0.27 | 0.32 ± 0.30 |
| MedNet-PVS | 0.20 ± 0.14[1,4,5] | 0.19 ± 0.14[1,4,5] | 0.53 ± 0.16[5] | 0.71 ± 0.25 |
| ADNI-PVS-Net | 0.60 ± 0.12 | 0.70 ± 0.13[1] | 0.74 ± 0.07 | 0.60 ± 0.19 |
| Rating 3 (n=14) | | | | |
| mcPVS-Net | 0.26 ± 0.09[2,4] | 0.33 ± 0.08[2,4,6] | 0.50 ± 0.07 | 0.72 ± 0.19 |
| nnU-Net T1 | 0.31 ± 0.11[2,4] | 0.38 ± 0.12[2,4] | 0.59 ± 0.05 | 0.52 ± 0.19 |
| nnU-Net T1+FLAIR | 0.30 ± 0.17 | 0.35 ± 0.17[2,4] | 0.55 ± 0.16 | 0.39 ± 0.22 |
| MedNet-PVS | 0.32 ± 0.10[2,4,6] | 0.31 ± 0.08[2,4,6] | 0.61 ± 0.06 | 0.71 ± 0.21 |
| ADNI-PVS-Net | 0.61 ± 0.15 | 0.69 ± 0.13 | 0.77 ± 0.06 | 0.56 ± 0.19 |
| Rating 4 (n=15) | | | | |
| mcPVS-Net | 0.33 ± 0.09[3,5] | 0.43 ± 0.11[3,5,6] | 0.52 ± 0.06 | 0.71 ± 0.11 |
| nnU-Net T1 | 0.36 ± 0.12[3,5] | 0.44 ± 0.15[3,5] | 0.59 ± 0.05 | 0.52 ± 0.19 |
| nnU-Net T1+FLAIR | 0.29 ± 0.17 | 0.35 ± 0.19[3,5] | 0.54 ± 0.17 | 0.34 ± 0.26 |
| MedNet-PVS | 0.40 ± 0.11[3,5,6] | 0.42 ± 0.12[3,5,6] | 0.62 ± 0.06[5] | 0.68 ± 0.12 |
| ADNI-PVS-Net | 0.60 ± 0.08 | 0.68 ± 0.07 | 0.75 ± 0.05 | 0.49 ± 0.15 |

Table 8B: Agreement with PVS severity – White matter

| Model | Weighted kappa | Percent agreement (%) |
|---|---|---|
| mcPVS-Net | 0.12 | 28.33 |
| nnU-Net T1 | 0.43 | 41.67 |
| nnU-Net T1+FLAIR | 0.47 | 46.67 |
| MedNet-PVS | 0.19 | 30.00 |
| ADNI-PVS-Net | 0.85 | 68.33 |

Table 9A: Performance by PVS severity categories — Basal ganglia

| Model | Overall | Object-level | Voxel-wise | Lesion-wise |
|---|---|---|---|---|
| Rating 1 (n=19) | | | | |
| mcPVS-Net | 0.08 ± 0.12[1,2,3] | 0.11 ± 0.12[1,2,3] | 0.38 ± 0.34 | 0.24 ± 0.27 |
| nnU-Net T1 | 0.09 ± 0.17[1,2,3] | 0.10 ± 0.13[2,3] | 0.29 ± 0.33 | 0.20 ± 0.22 |
| nnU-Net T1+FLAIR | 0.05 ± 0.12 | 0.05 ± 0.09 | 0.14 ± 0.24 | 0.08 ± 0.15 |
| MedNet-PVS | 0.09 ± 0.15[1,2,3] | 0.10 ± 0.10[1,2,3] | 0.34 ± 0.31 | 0.27 ± 0.28 |
| ADNI-PVS-Net | 0.22 ± 0.28[2,3] | 0.32 ± 0.38 | 0.32 ± 0.35[2,3] | 0.27 ± 0.34 |
| Rating 2 (n=12) | | | | |
| mcPVS-Net | 0.20 ± 0.12[1,5] | 0.28 ± 0.12[1,5] | 0.47 ± 0.19 | 0.46 ± 0.28 |
| nnU-Net T1 | 0.17 ± 0.16[1,5] | 0.20 ± 0.12[5] | 0.48 ± 0.26 | 0.27 ± 0.21 |
| nnU-Net T1+FLAIR | 0.15 ± 0.17 | 0.17 ± 0.14 | 0.37 ± 0.30 | 0.20 ± 0.20 |
| MedNet-PVS | 0.20 ± 0.13[1,5] | 0.25 ± 0.13[1,5] | 0.51 ± 0.18 | 0.49 ± 0.31 |
| ADNI-PVS-Net | 0.44 ± 0.24 | 0.57 ± 0.25 | 0.64 ± 0.23 | 0.47 ± 0.27 |
| Rating 3 (n=17) | | | | |
| mcPVS-Net | 0.21 ± 0.10[2,6] | 0.28 ± 0.13[2,6] | 0.49 ± 0.20 | 0.38 ± 0.25 |
| nnU-Net T1 | 0.23 ± 0.14[2,6] | 0.27 ± 0.12[2] | 0.52 ± 0.12 | 0.30 ± 0.21 |
| nnU-Net T1+FLAIR | 0.12 ± 0.15 | 0.15 ± 0.18 | 0.25 ± 0.26 | 0.11 ± 0.17 |
| MedNet-PVS | 0.24 ± 0.10[2,6] | 0.27 ± 0.09[2,6] | 0.50 ± 0.19 | 0.50 ± 0.27 |
| ADNI-PVS-Net | 0.49 ± 0.23[2] | 0.58 ± 0.24 | 0.67 ± 0.24[2] | 0.43 ± 0.23 |
| Rating 4 (n=12) | | | | |
| mcPVS-Net | 0.39 ± 0.08[3,5,6] | 0.48 ± 0.11[3,5,6] | 0.61 ± 0.07 | 0.43 ± 0.20 |
| nnU-Net T1 | 0.36 ± 0.10[3,5,6] | 0.38 ± 0.10[3,5] | 0.59 ± 0.06 | 0.29 ± 0.16 |
| nnU-Net T1+FLAIR | 0.14 ± 0.17 | 0.16 ± 0.17 | 0.33 ± 0.27 | 0.09 ± 0.14 |
| MedNet-PVS | 0.41 ± 0.10[3,5,6] | 0.45 ± 0.14[3,5,6] | 0.60 ± 0.10 | 0.58 ± 0.24 |
| ADNI-PVS-Net | 0.53 ± 0.18[3] | 0.58 ± 0.17 | 0.71 ± 0.10[3] | 0.36 ± 0.21 |

Table 9B: Agreement with PVS severity – Basal ganglia

| Model | Weighted kappa | Percent Agreement (%) |
|---|---|---|
| mcPVS-Net | 0.23 | 28.33 |
| nnU-Net T1 | 0.43 | 46.67 |
| nnU-Net T1+FLAIR | 0.11 | 35.00 |
| MedNet-PVS | 0.25 | 26.67 |
| ADNI-PVS-Net | 0.78 | 66.67 |

*Tables 8A, 9A: Values are mean ± SD. Superscript numbers denote a statistically significant pairwise difference between PVS severity ratings (Mann-Whitney U test, FDR-corrected across the 6 pairwise comparisons within each model/metric, $p < 0.05$).*
*Key: 1 = Rating 1 vs Rating 2; 2 = Rating 1 vs Rating 3; 3 = Rating 1 vs Rating 4; 4 = Rating 2 vs Rating 3; 5 = Rating 2 vs Rating 4; 6 = Rating 3 vs Rating 4*

Table 10A: Regression results — Whole Brain

| Model | WMH | Age | Sex | Aβ+/- | HT |
|---|---|---|---|---|---|
| PVS Count | | | | | |
| mcPVS-Net | $\beta_{exp}$ = 1.11, CI = [0.96, 1.28], p = 0.73 | $\beta_{exp}$ = 1.01, CI = [0.99, 1.02], p = 0.47 | $\beta_{exp}$ = 1.00, CI = [0.78, 1.28], p = 0.98 | $\beta_{exp}$ = 1.40, CI = [1.07, 1.83], p = 0.03 | $\beta_{exp}$ = 0.80, CI = [0.62, 1.04], p = 0.23 |
| nnU-Net T1 | $\beta_{exp}$ = 1.14, CI = [0.91, 1.41], p = 0.73 | $\beta_{exp}$ = 1.00, CI = [0.98, 1.02], p = 0.98 | $\beta_{exp}$ = 1.03, CI = [0.71, 1.51], p = 0.98 | $\beta_{exp}$ = 1.36, CI = [0.91, 2.05], p = 0.16 | $\beta_{exp}$ = 0.73, CI = [0.49, 1.08], p = 0.23 |
| nnU-Net T1+FLAIR | $\beta_{exp}$ = 1.07, CI = [0.80, 1.45], p = 0.73 | $\beta_{exp}$ = 0.99, CI = [0.97, 1.02], p = 0.98 | $\beta_{exp}$ = 1.16, CI = [0.69, 1.93], p = 0.98 | $\beta_{exp}$ = 1.14, CI = [0.66, 1.97], p = 0.64 | $\beta_{exp}$ = 0.82, CI = [0.48, 1.39], p = 0.46 |
| MedNet-PVS | $\beta_{exp}$ = 1.04, CI = [0.89, 1.22], p = 0.73 | $\beta_{exp}$ = 1.00, CI = [0.99, 1.02], p = 0.98 | $\beta_{exp}$ = 0.96, CI = [0.73, 1.27], p = 0.98 | $\beta_{exp}$ = 1.29, CI = [0.96, 1.73], p = 0.14 | $\beta_{exp}$ = 0.71, CI = [0.53, 0.95], p = 0.13 |
| ADNI-PVS-Net | $\beta_{exp}$ = 1.08, CI = [0.84, 1.40], p = 0.73 | $\beta_{exp}$ = 1.02, CI = [1.00, 1.04], p = 0.34 | $\beta_{exp}$ = 0.93, CI = [0.60, 1.44], p = 0.98 | $\beta_{exp}$ = 2.14, CI = [1.32, 3.46], p = 0.01 | $\beta_{exp}$ = 0.82, CI = [0.52, 1.30], p = 0.46 |
| Ground Truth | $\beta_{exp}$ = 0.95, CI = [0.70, 1.29], p = 0.73 | $\beta_{exp}$ = 1.02, CI = [1.00, 1.05], p = 0.33 | $\beta_{exp}$ = 0.94, CI = [0.57, 1.55], p = 0.98 | $\beta_{exp}$ = 2.46, CI = [1.44, 4.22], p = 0.01 | $\beta_{exp}$ = 0.75, CI = [0.45, 1.27], p = 0.43 |
| PVS Volume | | | | | |
| mcPVS-Net | $\beta_{exp}$ = 1.14, CI = [0.97, 1.34], p = 0.30 | $\beta_{exp}$ = 1.01, CI = [0.99, 1.03], p = 0.61 | $\beta_{exp}$ = 0.92, CI = [0.68, 1.25], p = 0.95 | $\beta_{exp}$ = 1.52, CI = [1.11, 2.09], p = 0.02 | $\beta_{exp}$ = 0.81, CI = [0.59, 1.09], p = 0.41 |

| | | | | | |
|---|---|---|---|---|---|
| nnU-Net T1 | $\beta_{exp}$ = 1.16, CI = [0.95, 1.41], p = 0.30 | $\beta_{exp}$ = 0.99, CI = [0.97, 1.02], p = 0.61 | $\beta_{exp}$ = 0.89, CI = [0.62, 1.30], p = 0.95 | $\beta_{exp}$ = 1.42, CI = [0.96, 2.10], p = 0.12 | $\beta_{exp}$ = 0.78, CI = [0.54, 1.14], p = 0.41 |
| nnU-Net T1+FLAIR | $\beta_{exp}$ = 0.99, CI = [0.74, 1.32], p = 0.94 | $\beta_{exp}$ = 0.99, CI = [0.96, 1.02], p = 0.61 | $\beta_{exp}$ = 1.02, CI = [0.60, 1.74], p = 0.95 | $\beta_{exp}$ = 1.25, CI = [0.71, 2.21], p = 0.44 | $\beta_{exp}$ = 0.88, CI = [0.51, 1.51], p = 0.69 |
| MedNet-PVS | $\beta_{exp}$ = 1.13, CI = [0.98, 1.31], p = 0.30 | $\beta_{exp}$ = 0.99, CI = [0.98, 1.01], p = 0.61 | $\beta_{exp}$ = 0.94, CI = [0.71, 1.24], p = 0.95 | $\beta_{exp}$ = 1.25, CI = [0.94, 1.67], p = 0.16 | $\beta_{exp}$ = 0.77, CI = [0.58, 1.01], p = 0.37 |
| ADNI-PVS-Net | $\beta_{exp}$ = 1.11, CI = [0.85, 1.46], p = 0.64 | $\beta_{exp}$ = 1.01, CI = [0.98, 1.04], p = 0.61 | $\beta_{exp}$ = 0.92, CI = [0.56, 1.52], p = 0.95 | $\beta_{exp}$ = 2.30, CI = [1.35, 3.92], p = 0.01 | $\beta_{exp}$ = 0.82, CI = [0.49, 1.37], p = 0.68 |
| Ground Truth | $\beta_{exp}$ = 0.91, CI = [0.66, 1.26], p = 0.70 | $\beta_{exp}$ = 1.02, CI = [0.98, 1.05], p = 0.61 | $\beta_{exp}$ = 0.98, CI = [0.54, 1.78], p = 0.95 | $\beta_{exp}$ = 2.72, CI = [1.45, 5.14], p = 0.01 | $\beta_{exp}$ = 0.88, CI = [0.48, 1.63], p = 0.69 |

Table 10B: Regression results — White Matter

| Model | WMH | Age | Sex | Aβ+/- | HT |
|---|---|---|---|---|---|
| PVS Count | | | | | |
| mcPVS-Net | $\beta_{exp}$ = 1.11, CI = [0.94, 1.30], p = 0.86 | $\beta_{exp}$ = 1.01, CI = [0.99, 1.02], p = 0.56 | $\beta_{exp}$ = 1.00, CI = [0.77, 1.32], p = 0.97 | $\beta_{exp}$ = 1.41, CI = [1.06, 1.90], p = 0.04 | $\beta_{exp}$ = 0.79, CI = [0.59, 1.05], p = 0.32 |
| nnU-Net T1 | $\beta_{exp}$ = 1.10, CI = [0.83, 1.47], p = 0.86 | $\beta_{exp}$ = 1.00, CI = [0.97, 1.02], p = 0.95 | $\beta_{exp}$ = 1.01, CI = [0.62, 1.65], p = 0.97 | $\beta_{exp}$ = 1.44, CI = [0.85, 2.44], p = 0.21 | $\beta_{exp}$ = 0.77, CI = [0.46, 1.28], p = 0.54 |
| nnU-Net T1+FLAIR | $\beta_{exp}$ = 0.98, CI = [0.63, 1.53], p = 0.94 | $\beta_{exp}$ = 0.99, CI = [0.96, 1.03], p = 0.95 | $\beta_{exp}$ = 1.06, CI = [0.51, 2.20], p = 0.97 | $\beta_{exp}$ = 1.26, CI = [0.57, 2.76], p = 0.57 | $\beta_{exp}$ = 0.90, CI = [0.42, 1.96], p = 0.80 |
| MedNet-PVS | $\beta_{exp}$ = 1.03, CI = [0.86, 1.24], p = 0.86 | $\beta_{exp}$ = 1.00, CI = [0.98, 1.02], p = 0.95 | $\beta_{exp}$ = 0.98, CI = [0.72, 1.34], p = 0.97 | $\beta_{exp}$ = 1.27, CI = [0.91, 1.78], p = 0.21 | $\beta_{exp}$ = 0.69, CI = [0.50, 0.96], p = 0.18 |
| ADNI-PVS-Net | $\beta_{exp}$ = 1.06, CI = [0.79, 1.44], p = 0.86 | $\beta_{exp}$ = 1.02, CI = [0.99, 1.04], p = 0.56 | $\beta_{exp}$ = 0.94, CI = [0.56, 1.58], p = 0.97 | $\beta_{exp}$ = 2.12, CI = [1.20, 3.74], p = 0.03 | $\beta_{exp}$ = 0.82, CI = [0.48, 1.39], p = 0.55 |
| Ground Truth | $\beta_{exp}$ = 0.91, CI = [0.64, 1.28], p = 0.86 | $\beta_{exp}$ = 1.02, CI = [1.00, 1.05], p = 0.45 | $\beta_{exp}$ = 0.95, CI = [0.55, 1.64], p = 0.97 | $\beta_{exp}$ = 2.48, CI = [1.36, 4.53], p = 0.02 | $\beta_{exp}$ = 0.77, CI = [0.44, 1.35], p = 0.54 |
| PVS Volume | | | | | |

| | | | | | |
|---|---|---|---|---|---|
| mcPVS-Net | $\beta_{exp}$ = 1.13, CI = [0.95, 1.33], p = 0.45 | $\beta_{exp}$ = 1.01, CI = [0.99, 1.03], p = 0.56 | $\beta_{exp}$ = 0.93, CI = [0.67, 1.27], p = 1.00 | $\beta_{exp}$ = 1.52, CI = [1.09, 2.13], p = 0.03 | $\beta_{exp}$ = 0.79, CI = [0.57, 1.09], p = 0.46 |
| nnU-Net T1 | $\beta_{exp}$ = 1.19, CI = [0.95, 1.50], p = 0.45 | $\beta_{exp}$ = 0.99, CI = [0.97, 1.02], p = 0.56 | $\beta_{exp}$ = 0.93, CI = [0.60, 1.43], p = 1.00 | $\beta_{exp}$ = 1.46, CI = [0.92, 2.31], p = 0.16 | $\beta_{exp}$ = 0.77, CI = [0.49, 1.19], p = 0.48 |
| nnU-Net T1+FLAIR | $\beta_{exp}$ = 0.97, CI = [0.70, 1.33], p = 0.84 | $\beta_{exp}$ = 0.99, CI = [0.95, 1.02], p = 0.56 | $\beta_{exp}$ = 1.00, CI = [0.55, 1.83], p = 1.00 | $\beta_{exp}$ = 1.36, CI = [0.72, 2.58], p = 0.35 | $\beta_{exp}$ = 0.95, CI = [0.52, 1.77], p = 0.88 |
| MedNet-PVS | $\beta_{exp}$ = 1.11, CI = [0.93, 1.31], p = 0.45 | $\beta_{exp}$ = 0.99, CI = [0.97, 1.01], p = 0.56 | $\beta_{exp}$ = 0.97, CI = [0.70, 1.34], p = 1.00 | $\beta_{exp}$ = 1.24, CI = [0.88, 1.74], p = 0.27 | $\beta_{exp}$ = 0.75, CI = [0.54, 1.04], p = 0.46 |
| ADNI-PVS-Net | $\beta_{exp}$ = 1.08, CI = [0.81, 1.45], p = 0.70 | $\beta_{exp}$ = 1.01, CI = [0.98, 1.04], p = 0.66 | $\beta_{exp}$ = 0.98, CI = [0.57, 1.71], p = 1.00 | $\beta_{exp}$ = 2.27, CI = [1.27, 4.06], p = 0.02 | $\beta_{exp}$ = 0.79, CI = [0.45, 1.39], p = 0.62 |
| Ground Truth | $\beta_{exp}$ = 0.84, CI = [0.60, 1.17], p = 0.45 | $\beta_{exp}$ = 1.01, CI = [0.98, 1.05], p = 0.56 | $\beta_{exp}$ = 1.06, CI = [0.56, 1.98], p = 1.00 | $\beta_{exp}$ = 2.60, CI = [1.33, 5.07], p = 0.02 | $\beta_{exp}$ = 0.87, CI = [0.46, 1.66], p = 0.81 |

Table 10C: Regression results — Basal Ganglia

| Model | WMH | Age | Sex | Aβ+/- | HT |
|---|---|---|---|---|---|
| PVS Count | | | | | |
| mcPVS-Net | $\beta_{exp}$ = 1.16, CI = [1.01, 1.34], p = 0.13 | $\beta_{exp}$ = 1.01, CI = [0.99, 1.03], p = 0.57 | $\beta_{exp}$ = 0.90, CI = [0.69, 1.17], p = 0.58 | $\beta_{exp}$ = 1.27, CI = [0.96, 1.69], p = 0.18 | $\beta_{exp}$ = 0.93, CI = [0.70, 1.22], p = 0.67 |
| nnU-Net T1 | $\beta_{exp}$ = 0.97, CI = [0.83, 1.14], p = 0.74 | $\beta_{exp}$ = 1.00, CI = [0.98, 1.02], p = 0.98 | $\beta_{exp}$ = 0.92, CI = [0.69, 1.23], p = 0.58 | $\beta_{exp}$ = 1.15, CI = [0.84, 1.56], p = 0.57 | $\beta_{exp}$ = 0.73, CI = [0.54, 0.99], p = 0.18 |
| nnU-Net T1+FLAIR | $\beta_{exp}$ = 0.79, CI = [0.54, 1.15], p = 0.26 | $\beta_{exp}$ = 0.98, CI = [0.94, 1.01], p = 0.57 | $\beta_{exp}$ = 0.68, CI = [0.34, 1.36], p = 0.56 | $\beta_{exp}$ = 0.94, CI = [0.45, 1.97], p = 0.87 | $\beta_{exp}$ = 0.54, CI = [0.26, 1.12], p = 0.20 |
| MedNet-PVS | $\beta_{exp}$ = 1.12, CI = [0.97, 1.29], p = 0.26 | $\beta_{exp}$ = 1.01, CI = [0.99, 1.02], p = 0.57 | $\beta_{exp}$ = 0.92, CI = [0.70, 1.21], p = 0.58 | $\beta_{exp}$ = 1.09, CI = [0.82, 1.45], p = 0.68 | $\beta_{exp}$ = 0.76, CI = [0.57, 1.01], p = 0.18 |
| ADNI-PVS-Net | $\beta_{exp}$ = 1.37, CI = [1.01, 1.85], p = 0.13 | $\beta_{exp}$ = 1.01, CI = [0.98, 1.05], p = 0.57 | $\beta_{exp}$ = 0.62, CI = [0.36, 1.07], p = 0.52 | $\beta_{exp}$ = 1.96, CI = [1.11, 3.48], p = 0.06 | $\beta_{exp}$ = 0.88, CI = [0.49, 1.59], p = 0.67 |
| Ground Truth | $\beta_{exp}$ = 1.23, CI = [0.90, 1.68], p = 0.26 | $\beta_{exp}$ = 1.01, CI = [0.98, 1.05], p = 0.57 | $\beta_{exp}$ = 0.72, CI = [0.41, 1.28], p = 0.56 | $\beta_{exp}$ = 2.25, CI = [1.25, 4.05], p = 0.04 | $\beta_{exp}$ = 0.76, CI = [0.41, 1.39], p = 0.56 |
| PVS Volume | | | | | |

| | | | | | |
|---|---|---|---|---|---|
| mcPVS-Net | $\beta_{exp}$ = 1.25, CI = [1.03, 1.53], p = 0.15 | $\beta_{exp}$ = 1.01, CI = [0.99, 1.04], p = 0.43 | $\beta_{exp}$ = 0.85, CI = [0.58, 1.23], p = 0.38 | $\beta_{exp}$ = 1.25, CI = [0.84, 1.85], p = 0.54 | $\beta_{exp}$ = 0.93, CI = [0.64, 1.35], p = 0.70 |
| nnU-Net T1 | $\beta_{exp}$ = 0.95, CI = [0.80, 1.13], p = 0.57 | $\beta_{exp}$ = 1.00, CI = [0.99, 1.02], p = 0.66 | $\beta_{exp}$ = 0.76, CI = [0.55, 1.04], p = 0.21 | $\beta_{exp}$ = 1.08, CI = [0.77, 1.51], p = 0.95 | $\beta_{exp}$ = 0.77, CI = [0.56, 1.07], p = 0.37 |
| nnU-Net T1+FLAIR | $\beta_{exp}$ = 0.82, CI = [0.60, 1.11], p = 0.29 | $\beta_{exp}$ = 0.98, CI = [0.94, 1.01], p = 0.43 | $\beta_{exp}$ = 0.50, CI = [0.28, 0.88], p = 0.10 | $\beta_{exp}$ = 0.96, CI = [0.52, 1.77], p = 0.95 | $\beta_{exp}$ = 0.48, CI = [0.27, 0.87], p = 0.09 |
| MedNet-PVS | $\beta_{exp}$ = 1.21, CI = [0.98, 1.49], p = 0.24 | $\beta_{exp}$ = 1.02, CI = [1.00, 1.04], p = 0.43 | $\beta_{exp}$ = 0.82, CI = [0.55, 1.23], p = 0.38 | $\beta_{exp}$ = 0.99, CI = [0.65, 1.50], p = 0.95 | $\beta_{exp}$ = 0.80, CI = [0.53, 1.20], p = 0.55 |
| ADNI-PVS-Net | $\beta_{exp}$ = 1.31, CI = [0.89, 1.94], p = 0.29 | $\beta_{exp}$ = 1.01, CI = [0.97, 1.06], p = 0.64 | $\beta_{exp}$ = 0.54, CI = [0.26, 1.14], p = 0.21 | $\beta_{exp}$ = 1.66, CI = [0.76, 3.63], p = 0.54 | $\beta_{exp}$ = 0.83, CI = [0.39, 1.75], p = 0.70 |
| Ground Truth | $\beta_{exp}$ = 1.24, CI = [0.83, 1.85], p = 0.36 | $\beta_{exp}$ = 1.02, CI = [0.98, 1.07], p = 0.46 | $\beta_{exp}$ = 0.60, CI = [0.28, 1.29], p = 0.29 | $\beta_{exp}$ = 2.25, CI = [1.01, 5.01], p = 0.29 | $\beta_{exp}$ = 0.71, CI = [0.33, 1.54], p = 0.58 |

*Tables 10A-C: Regression results for whole brain, white matter (WM) and basal ganglia (BG) are shown in these tables. β-coefficients and confidence intervals are reported as the exponent of the original value for interpretability since regressions are modeled either using a Negative Binomial (PVS count) or Gamma GLM with a log-link function (PVS volume). Aβ status was modeled with Aβ− as the reference group, so values > 1 indicate higher PVS burden in Aβ+ participants.*
